# Sex differences in associations between adversity and biological ageing

**DOI:** 10.1101/2025.08.28.25334645

**Authors:** Julian Mutz, Luca Di Benedetto, Thole H Hoppen, Nexhmedin Morina, Monica Aas

## Abstract

**Background:** Adverse events across the life course have been linked to older biological ageing profiles. Whether these associations differ between males and females, and whether such differences depend on adversity occurring in childhood, adulthood or both periods, remains unclear.

**Methods:** In 153,557 UK Biobank participants aged 40-69 years, we assessed associations of childhood and/or adulthood adversity with metabolomic ageing, frailty, telomere length and grip strength. Sex differences were evaluated using stratified analyses and sex-by-adversity interaction tests.

**Results:** Exposure to adversity in childhood and/or adulthood was reported by 64.6% of males and 69.6% of females. Childhood adversity was more strongly associated with multiple ageing markers in females, including a metabolite-predicted age exceeding chronological age, greater frailty, shorter telomeres and weaker grip strength. Adulthood adversity was more strongly associated with certain ageing markers in males, particularly greater frailty and weaker grip strength. This divergence in sex-specific associations between childhood and adulthood exposure was consistent across several markers, with statistically significant sex-by-adversity interactions for frailty and grip strength.

**Conclusions:** In this large, population-based sample, the timing of adversity, distinguishing childhood from adulthood, shaped whether females or males showed stronger associations with biological ageing markers. These findings suggest that sex differences in biological ageing profiles may partly reflect distinct sensitive periods of vulnerability, highlighting the importance of considering both sex and timing of exposure to adversity when examining links between adversity and biological ageing.

## Introduction

Adversity in childhood and adulthood has been linked to a wide array of molecular, clinical and functional markers of biological ageing (Aas et al., 2025; Bourassa & Sbarra, 2024). Childhood adversity, in particular, may contribute to various disease processes and increase the risk of excess morbidity and mortality (Alley, Gassen, & Slavich, 2025; Bourassa et al., 2023). Proposed mechanisms include telomere attrition, increased oxidative stress and heightened systemic inflammation, all of which are linked to accelerated ageing (Bourassa et al., 2023; Polsky, Rentscher, & Carroll, 2022; Yegorov, Poznyak, Nikiforov, Sobenin, & Orekhov, 2020). Despite robust evidence linking adversity to ageing markers, few large, population-based studies have investigated whether these associations vary by sex and across different ageing markers. Clarifying such differences could shed light on sex-specific disparities in health, life expectancy and resilience (Austad & Bartke, 2015; DuMont, Widom, & Czaja, 2007), and inform tailored prevention and intervention strategies.

In our prior study, individuals exposed to adversity in both childhood and adulthood had a metabolite-predicted age (MileAge) exceeding their chronological age, greater frailty and weaker grip strength; those exposed to childhood adversity, particularly abuse, also had shorter telomeres (Aas et al., 2025). However, the role of biological sex in moderating these associations remains unclear. Exposure to adversity and its health consequences often differ between males and females due to a combination of biological, behavioural and social factors. Females report a higher overall incidence of adversity (Merrick, Ford, Ports, & Guinn, 2018), particularly childhood sexual abuse, while other adversities such as physical abuse or combat are more common in males (Aas et al., 2016; Fisher et al., 2009; Mersky, Choi, Plummer Lee, & Janczewski, 2021). Prior research has also identified sex differences in the prevalence of depression and anxiety associated with childhood adversity (Zhu et al., 2025). Amongst adversity-exposed individuals, females tend to experience earlier illness onset, more depressive episodes, more suicide attempts and more rapid mood cycling than males (Etain et al., 2013; Fisher et al., 2009).

Although females report a higher incidence of adversity overall, they may benefit from the protective hormonal effects of oestrogen, which modulates stress responses, reduces inflammation and supports neuroplasticity (Galea, Frick, Hampson, Sohrabji, & Choleris, 2017). These effects may confer resilience, potentially mitigating the negative impact of adversity on biological ageing. Conversely, relatively lower oestrogen levels in males may contribute to greater vulnerability to stress-induced biological ageing (Gurvich, Hoy, Thomas, & Kulkarni, 2018; Horstman, Dillon, Urban, & Sheffield-Moore, 2012) and result in divergent psychiatric and cognitive outcomes (Wei et al., 2014). However, some evidence suggests that higher testosterone levels in males may buffer stress responses by blunting cortisol release, thereby reducing hypothalamic-pituitary adrenal axis activation (Bale & Epperson, 2015). Social and behavioural factors, such as differential coping strategies and the availability of social support networks, may further shape sex-specific responses to adversity (Kim, Song, & Lee, 2017; Kundakovic, Lim, Gudsnuk, & Champagne, 2013). Consistent with this resilience hypothesis, these factors appear to confer an advantage to females, who have a longer life expectancy than males (Zarulli et al., 2018).

Despite a growing interest in the link between adversity and biological ageing, prior research examining sex differences in these associations has limitations such as reliance on small samples (Hägg & Jylhävä, 2021), focus on only a single ageing marker (e.g., telomere length (Ridout, Ridout, Price, Sen, & Tyrka, 2016)) and lack of integration with biobank data (Yu et al., 2024). Addressing these gaps through larger, multi-marker studies will substantially strengthen the validity and generalisability of findings. It also remains unknown whether sex differences in the associations between adversity and ageing markers vary according to when in the life course adversity occurs, for example, during sensitive developmental periods in childhood versus adulthood.

In this study, we used data from the UK Biobank to investigate sex differences in associations between childhood and adulthood adversity and molecular, clinical and functional markers of biological ageing. Specifically, we examined associations with metabolomic ageing, frailty, telomere length and grip strength. Extending prior research, we performed both sex-stratified analyses and formally tested sex-by-adversity interactions in a large, well-characterised, population-based cohort. We examined both childhood and adulthood adversity to determine whether the timing of exposure modifies the direction or magnitude of sex differences in biological ageing profiles. Given emerging evidence that relational trauma, i.e. trauma occurring within the contexts of close relationships, may be especially harmful (Anders, Shallcross, & Frazier, 2012), our analyses primarily focused on relational forms of adversity in childhood and/or adulthood. Based on prior findings and the proposed protective role of oestrogen, we hypothesised that although females would report more adversity overall, males would show stronger associations between adversity exposure and markers of biological ageing. We also explored the possibility that any sex differences might vary by timing of exposure, with childhood adversity potentially producing distinct patterns from adulthood adversity.

## Methods

### Study population

The UK Biobank recruited over 500,000 individuals (37-73 years) from England, Scotland and Wales between 2006 and 2010. At baseline, participants completed health and sociodemographic questionnaires, underwent physical examinations and provided biological samples.

### Adverse events

Exposure to adversity was assessed in 2016-2017 using the online Mental Health Questionnaire (MHQ) (Davis et al., 2020).

Childhood adverse events were assessed using the Childhood Trauma Screener (Glaesmer et al., 2013), a short version of the Childhood Trauma Questionnaire (Bernstein & Fink, 1998). It comprises five items, with responses ranging from “never true” to “very often true”: “When I was growing up,…1) I felt loved [emotional neglect]; 2) people in my family hit me so hard that it left me with bruises or marks [physical abuse]; 3) I felt that someone in my family hated me [emotional abuse]; 4) someone molested me sexually [sexual abuse]; and 5) there was someone to take me to the doctor if I needed it [physical neglect; reverse coded].” Individuals were classified as having experienced adversity in childhood based on the following cut-offs: physical abuse, emotional abuse or sexual abuse (at least “rarely true”); emotional neglect or physical neglect (less than “often”; reverse coded). We then derived a sum score (ranging from zero to five), reflecting the overall childhood adversity burden.

Adulthood adverse events were assessed using questions adapted from the 2010/2011 British Crime Survey to identify victims of crime and domestic violence (Khalifeh, Oram, Trevillion, Johnson, & Howard, 2015). Five questions were asked, with responses ranging from “never true” to “very often true”: “Since I was sixteen,…1) I have been in a confiding relationship [emotional neglect]; 2) a partner or ex-partner repeatedly belittled me to the extent that I felt worthless [emotional abuse]; 3) a partner or ex-partner deliberately hit me or used violence in any other way [physical abuse]; 4) a partner or ex-partner sexually interfered with me, or forced me to have sex against my wishes [sexual abuse]; 5) there was money to pay the rent or mortgage when I needed it [economic hardship; reverse coded].” Individuals were classified as having experienced adulthood adversity based on the following cut-offs: emotional abuse, sexual abuse or physical violence (at least “rarely true”); emotional neglect (“sometimes true” or less; reverse coded); economic hardship (“often” or less; reverse coded). We then derived a sum score (ranging from zero to five), reflecting overall adulthood adversity burden.

### Metabolomic age (MileAge) delta

Nuclear magnetic resonance spectroscopy-derived metabolomic biomarkers were quantified in non-fasting plasma samples using the Nightingale Health platform, which ascertains 249 biomarkers (168 in absolute concentrations and 81 derived ratios) (Würtz et al., 2017).

Technical variation was removed using the ‘ukbnmr’ R package (algorithm v2) (Ritchie et al., 2023). In a prior study (Mutz, Iniesta, & Lewis, 2024), we developed a metabolomic ageing clock using a Cubist rule-based regression model. Individual-level age predictions were aggregated from the outer loop of the nested cross-validation to avoid potential overfitting. Metabolomic age (MileAge) delta represents the difference between metabolite-predicted and chronological age, with positive values indicating an older biological age profile (Mutz et al., 2024).

### Metabolomic mortality profile score

In a previous study (Zhang et al., 2024), we developed a metabolomic mortality profile score. Complementing the 249 biomarkers provided by the Nightingale Health platform, 76 additional lipid, cholesterol and fatty acid ratios were derived (Ritchie et al., 2023), resulting in a total of 325 biomarkers. A Least Absolute Shrinkage and Selection Operator (LASSO) Cox proportional hazards model predicting mortality was developed in English and Welsh participants (*N* = 234,553). We derived a mortality profile score in Scottish participants (*N* = 15,788) as the linear combination of the 54 biomarkers with non-zero coefficients in the LASSO Cox model weighted by their log hazard ratio for mortality. Higher scores indicate an elevated mortality risk.

### Frailty index

A frailty index was derived from health deficits reported via touch-screen questionnaires or during nurse-led interviews that met the following criteria: indicators of poor health, more prevalent in older individuals, neither rare nor universal, covering multiple areas of functioning and available for ≥ 80% of participants (Williams, Jylhävä, Pedersen, & Hägg, 2019). The 49 items that met those criteria included cardiometabolic, cranial, immunological, musculoskeletal, respiratory and sensory traits, well-being, infirmity, cancer and pain.

Categorical variables were dichotomised (deficit absent = zero; deficit present = one) and ordinal variables were mapped onto a score between zero and one. The sum of deficits was divided by the number of possible deficits, resulting in frailty index scores between zero and one, with higher scores indicating greater frailty (Mutz, Choudhury, Zhao, & Dregan, 2022). Participants with missing data for ≥ 10/49 variables were excluded (Williams et al., 2019).

### Telomere length

Leukocyte telomere length was measured using a quantitative polymerase chain reaction (qPCR) assay that expresses telomere length as the ratio of the telomere repeat copy number (T) relative to a single-copy gene (S) encoding haemoglobin subunit beta (Codd et al., 2022). The T/S ratio is proportional to average telomere length (Lai, Wright, & Shay, 2018). Measurements were adjusted for operational and technical parameters (PCR machine, staff member, enzyme batch, primer batch, temperature, humidity, primer batch × PCR machine, primer batch × staff member, A260/A280 ratio of the DNA sample and A260/A280 ratio squared), log*_e_* transformed and *Z*-standardised.

### Grip strength

Grip strength in whole kilogram force units was measured using a Jamar J00105 hydraulic hand dynamometer (measurement range 0-90 kg) for both hands. We used the maximal grip strength of the participant’s self-reported dominant hand. If no data on handedness were available, we used the highest value.

### Covariates

Potential confounders included chronological age, highest educational/professional qualification, gross annual household income, ethnicity and neighbourhood deprivation (assessed via the Townsend deprivation index).

### Exclusion criteria

Individuals with missing data or who responded “prefer not to answer” to any questionnaire items on adversity were excluded from the derived variables (binary phenotypes and sum scores). For item-specific analyses, only those with missing data or with a “prefer not to answer” response for the specific item were excluded. For categorical covariates, “do not know” responses, “prefer not to answer” responses and missing data were coded as a missing data level. For the analyses of MileAge delta, we applied exclusions as per the original study (Mutz et al., 2024): females with possible pregnancy, as metabolite profiles differ during pregnancy; individuals with discordant genetic and self-reported sex; and individuals with missing or outlier metabolite values (4× the interquartile range from the median).

### Statistical analyses

Data processing, analyses and visualisations were performed in R (version 4.3.0). Sample characteristics were summarised using means and standard deviations or counts and percentages. To explore potential sex differences, we estimated sex-stratified associations and tested sex-by-adversity interactions by adding cross-product terms to regression models.

Associations between adverse events (exposures) and MileAge delta, the metabolomic mortality profile score, the frailty index, telomere length and grip strength (outcomes) were estimated using ordinary least squares regression. All outcomes were scaled to have a mean equal to zero and a standard deviation of one, allowing for direct comparison of the association estimates. Telomere length and grip strength were reverse coded prior to analysis. First, we examined sum scores of adverse childhood events and of adulthood events. Second, we examined associations with the cross-classification of childhood and adulthood adversity (childhood and adulthood; childhood only; adulthood only; neither), derived from binary definitions of childhood and adulthood adversity. For each outcome, we fitted a model adjusted for chronological age (Model 1) and a fully adjusted model also including education, income, ethnicity and deprivation (Model 2). *P*-values were adjusted for multiple testing using the Benjamini-Hochberg correction method, with a two-tailed test and a false discovery rate of 5%. To investigate whether specific types of adversity were associated with markers of biological ageing, we further examined associations for each individual item included in the questionnaires. We applied the same modelling strategy reported above.

## Results

### Childhood adversity

Amongst 153,557 individuals across sexes, 41.0% of males (27,488 out of 67,085) and 41.1% of females (35,578 out of 86,472) reported exposure to adverse events during childhood (Table S1). Analytical sample sizes are reported in Table S2; those include individuals with/without adulthood adversity. After full covariate adjustment, childhood adversity was associated with a metabolite-predicted age exceeding chronological age (*β* = 0.019, 95% CI 0.009-0.028, *p* < 0.001), shorter telomeres (*β* = 0.014, 95% CI 0.008-0.020, *p* < 0.001) and weaker grip strength (*β* = 0.010, 95% CI 0.007-0.013, *p* < 0.001) in females but not in males (Figure 1A; Table 1). Formal sex-by-adversity interaction tests, however, were not statistically significant (*p* _interaction_ > 0.063) (Table S3).

**Figure 1.**
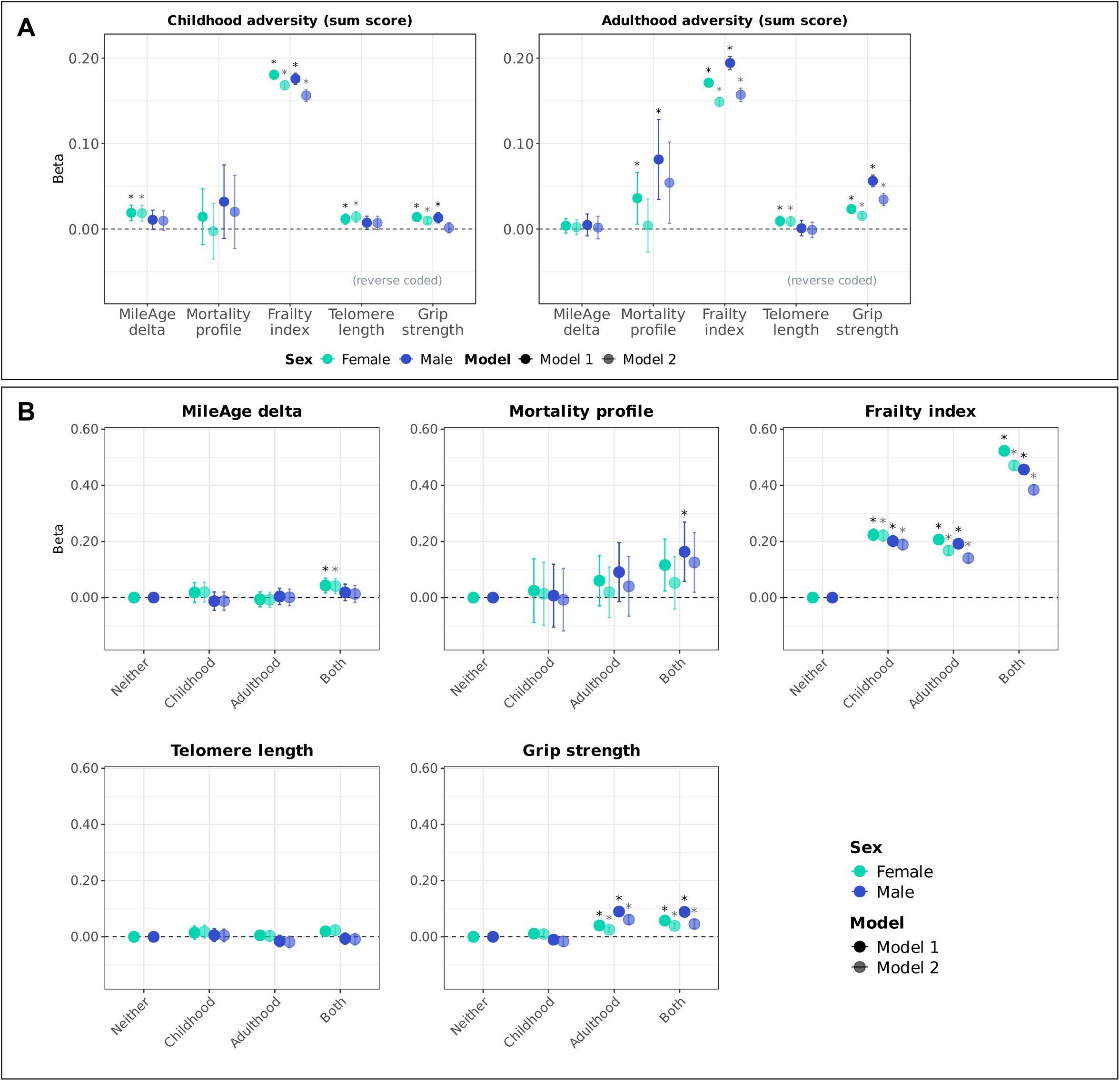
(A) Ageing markers and adversity (sum scores). Sex-stratified associations between adversity (adverse/traumatic events in childhood and in adulthood) and ageing markers (MileAge delta, frailty index, telomere length [reverse coded] and grip strength [reverse coded]). Asterisks indicate statistically significant associations, after correcting *p*-values for multiple testing using the Benjamini– Hochberg procedure (across childhood and adulthood exposures, ageing markers and models, separately for each sex). **(B) Ageing markers and adversity (cross-classification childhood and adulthood).** Sex-stratified associations between adversity (adverse/traumatic events in childhood and/or adulthood) and ageing markers (MileAge delta, frailty index, telomere length [reverse coded] and grip strength [reverse coded]). Asterisks indicate statistically significant associations, after correcting *p*-values for multiple testing using the Benjamini–Hochberg procedure (across exposure levels, ageing markers and models, separately for each sex). **(A-B)** Estimates shown are ordinary least squares regression beta coefficients and 95% confidence intervals. Model 1–adjusted for chronological age; Model 2–adjusted for chronological age, ethnicity, highest educational/professional qualification, annual gross household income and neighbourhood deprivation. Sample sizes reported in Tables S1, S4 and S6.

**Table 1.** Sex-stratified associations between adverse events and ageing markers.

| Females |  | Model 1 (adj. age) |  |  | Model 2 (full adjustment) |  |  |  |
| --- | --- | --- | --- | --- | --- | --- | --- | --- |
| Childhood | $\beta$ | 95% CI | | $p$ | $\beta$ | 95% CI | | $p$ |
| MileAge delta | 0.019 | 0.010 | 0.028 | <0.001 | 0.019 | 0.009 | 0.028 | <0.001 |
| Metabolomic profile | 0.014 | -0.018 | 0.047 | 0.456 | -0.003 | -0.035 | 0.030 | 0.880 |
| Frailty index | 0.180 | 0.175 | 0.186 | <0.001 | 0.168 | 0.163 | 0.174 | <0.001 |
| Telomere length | 0.012 | 0.006 | 0.018 | <0.001 | 0.014 | 0.008 | 0.020 | <0.001 |
| Grip strength | 0.014 | 0.011 | 0.017 | <0.001 | 0.010 | 0.007 | 0.013 | <0.001 |
| Adulthood |  |  |  |  |  |  |  |  |
| MileAge delta | 0.004 | -0.005 | 0.012 | 0.456 | 0.002 | -0.007 | 0.011 | 0.700 |
| Metabolomic profile | 0.036 | 0.006 | 0.066 | 0.027 | 0.004 | -0.027 | 0.035 | 0.834 |
| Frailty index | 0.171 | 0.166 | 0.176 | <0.001 | 0.149 | 0.144 | 0.154 | <0.001 |
| Telomere length | 0.009 | 0.004 | 0.015 | 0.002 | 0.009 | 0.003 | 0.015 | 0.004 |
| Grip strength | 0.023 | 0.020 | 0.026 | <0.001 | 0.015 | 0.012 | 0.019 | <0.001 |
| Males |  |  |  |  |  |  |  |  |
| Childhood |  |  |  |  |  |  |  |  |
| MileAge delta | 0.011 | -0.001 | 0.022 | 0.121 | 0.010 | -0.002 | 0.021 | 0.153 |
| Metabolomic profile | 0.032 | -0.011 | 0.075 | 0.206 | 0.020 | -0.023 | 0.063 | 0.478 |
| Frailty index | 0.176 | 0.169 | 0.182 | <0.001 | 0.156 | 0.149 | 0.163 | <0.001 |
| Telomere length | 0.007 | 0.000 | 0.015 | 0.121 | 0.007 | -0.001 | 0.015 | 0.131 |
| Grip strength | 0.013 | 0.007 | 0.019 | <0.001 | 0.001 | -0.005 | 0.007 | 0.745 |
| Adulthood |  |  |  |  |  |  |  |  |
| MileAge delta | 0.005 | -0.008 | 0.018 | 0.584 | 0.002 | -0.012 | 0.015 | 0.847 |
| Metabolomic profile | 0.081 | 0.035 | 0.128 | 0.002 | 0.054 | 0.007 | 0.102 | 0.057 |
| Frailty index | 0.194 | 0.186 | 0.202 | <0.001 | 0.157 | 0.149 | 0.165 | <0.001 |
| Telomere length | 0.001 | -0.008 | 0.010 | 0.847 | -0.001 | -0.010 | 0.008 | 0.847 |
| Grip strength | 0.056 | 0.050 | 0.063 | <0.001 | 0.034 | 0.028 | 0.041 | <0.001 |

*Note:* CI = confidence interval. Model 1—adjusted for chronological age; Model 2—adjusted for chronological age, ethnicity, highest educational/professional qualification, annual gross household income and neighbourhood deprivation. *P*-values shown are corrected for multiple testing using the Benjamini–Hochberg procedure (across childhood and adulthood exposures, ageing markers and models, separately for each sex). Sample sizes reported in Tables S1 and S4.

### Adulthood adversity

Amongst 150,848 individuals across sexes, 48.2% (31,830 out of 66,019) and 57.8% of females (49,065 out of 84,829) reported exposure to adverse events during adulthood (Table S4). Analytical sample sizes are reported in Table S5; those include individuals with/without childhood adversity. Adulthood adversity was associated with shorter telomeres in females (*β* = 0.009, 95% CI 0.003-0.015, *p* = 0.004) but not in males (*β* = −0.001, 95% CI −0.010 to 0.008, *p* = 0.847) (Figure 1A; Table 1). However, the sex-by-adversity interaction test was not statistically significant (*p* _interaction_ = 0.063) (Table S3). Associations between adulthood adversity and elevated metabolomic mortality profile scores, greater frailty and weaker grip strength were stronger in males than in females. Formal interaction tests confirmed stronger associations in males for frailty (*β* _interaction_ = 0.01, 95% CI 0.00-0.02, *p* = 0.017) and grip strength (*β* _interaction_ = 0.03, 95% CI 0.02-0.03, *p* < 0.001).

### Cross-classification between childhood and adulthood adversity

Amongst 147,958 individuals across sexes, 24.3% of males (15,852 out of 65,107) and 29.2% of females (24,234 out of 82,851) reported exposure to adverse events during both childhood and adulthood. Exposure to adversity in childhood and/or adulthood was reported by 64.6% of males and 69.6% of females. Analytical sample sizes are reported in Table S6. Adversity in both childhood and adulthood was associated with a metabolite-predicted age exceeding chronological age in females (*β* = 0.041, 95% CI 0.014-0.068, *p* = 0.009) but not in males (*β* = 0.013, 95% CI −0.017 to 0.043, *p* = 0.708) (Figure 1B; Table 2). However, the sex-by-adversity interaction test was not statistically significant (*β* _interaction_ = 0.00, 95% CI −0.04 to 0.04, *p* = 0.894) (Table S7). In contrast, having experienced adversity in both childhood and adulthood was associated with higher metabolomic mortality profile scores only in males (age-adjusted *β* = 0.163, 0.058-0.269, *p* = 0.014), though not after full adjustment (*β* = 0.125, 95% CI 0.019-0.232, *p* = 0.063). There was no evidence of interaction (*β* _interaction_ = 0.05, 95% CI −0.09 to 0.19, *p* = 0.723). Associations between adversity and frailty were generally stronger in females, with a statistically significant interaction observed for adversity in both childhood and adulthood (*β* _females_ = 0.471, 95% CI 0.456-0.487, *p* < 0.001; *β* _males_ = 0.384, 0.366-0.401, *p* < 0.001; *β* _interaction_ = −0.08, 95% CI −0.10 to −0.06, *p* < 0.001). Although associations between adversity in both childhood and adulthood and telomeres were neither statistically significant in females (*β* = 0.024, 95% CI 0.005-0.042, *p* = 0.063) nor in males (*β* = −0.009, 95% CI −0.029 to 0.012, *p* = 0.658), the interaction test was statistically significant (*β* _interaction_ = −0.04, 95% CI −0.07 to −0.01, *p* = 0.024). The association between adversity in adulthood only and grip strength was stronger in males (*β* = 0.061, 95% CI 0.046-0.077, *p* < 0.001) than in females (*β* = 0.026, 95% CI 0.016-0.035, *p* < 0.001), with a statistically significant interaction (*β* _interaction_ = 0.05, 95% CI 0.03-0.06, *p* < 0.001). The association between adversity in both childhood and adulthood and grip strength was modestly stronger in males (*β* = 0.046, 0.030-0.061, *p* < 0.001) than in females (*β* = 0.039, 95% CI 0.029-0.049, *p* < 0.001), but the interaction test did not reach statistical significance (*β* _interaction_ = 0.02, 95% CI 0.00-0.04, *p* = 0.051).

**Table 2.**
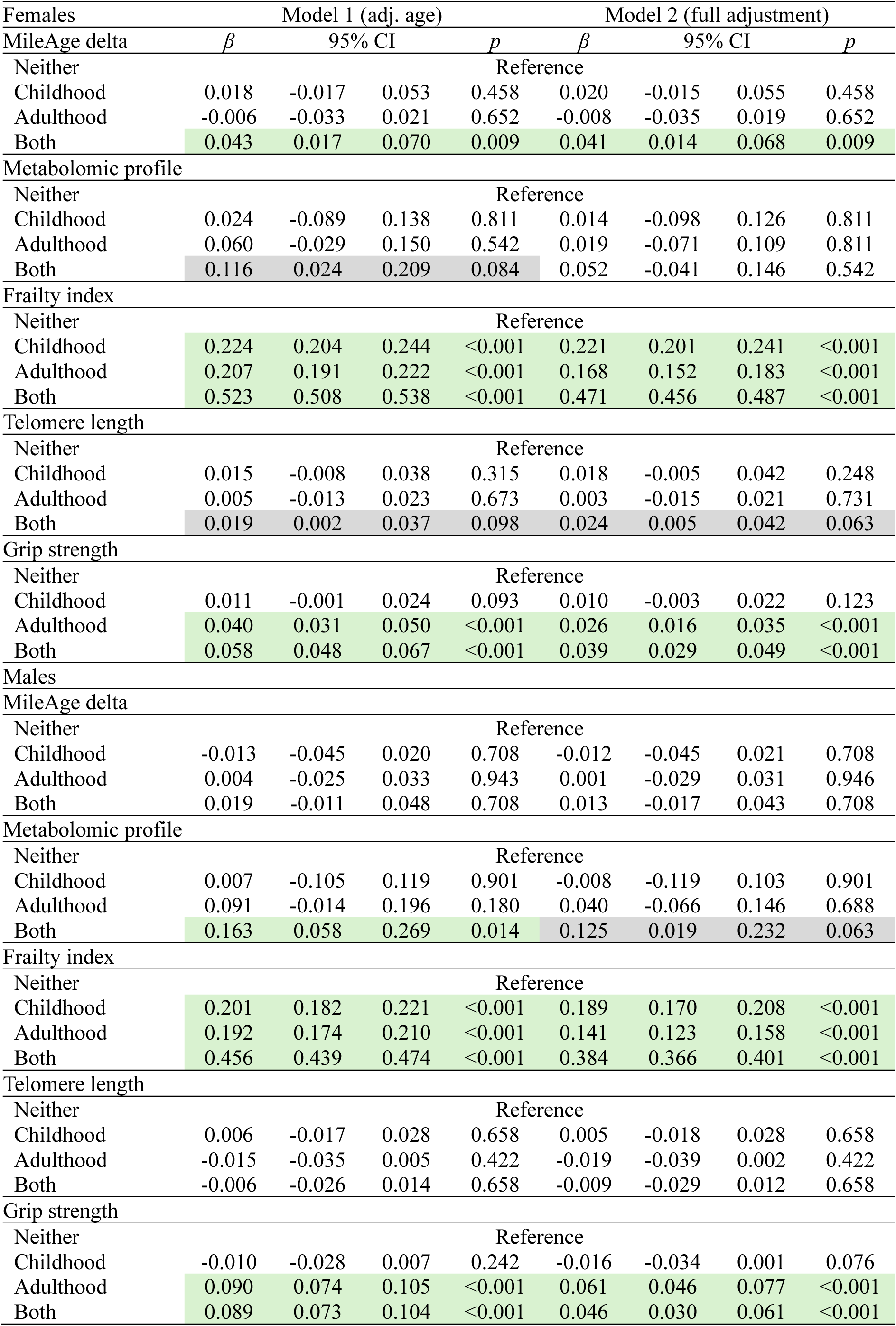
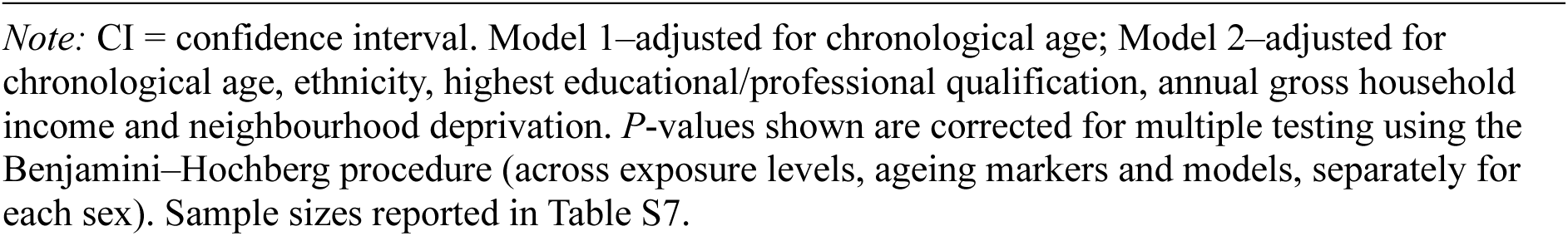
Sex-stratified associations between adverse events and ageing markers.

| Females |  | Model 1 (adj. age) |  |  | Model 2 (full adjustment) |  |  |  |
| --- | --- | --- | --- | --- | --- | --- | --- | --- |
| MileAge delta | $\beta$ | 95% CI | | $p$ | $\beta$ | 95% CI | | $p$ |
| Neither |  |  |  | Reference |  |  |  |  |
| Childhood | 0.018 | -0.017 | 0.053 | 0.458 | 0.020 | -0.015 | 0.055 | 0.458 |
| Adulthood | -0.006 | -0.033 | 0.021 | 0.652 | -0.008 | -0.035 | 0.019 | 0.652 |
| Both | 0.043 | 0.017 | 0.070 | 0.009 | 0.041 | 0.014 | 0.068 | 0.009 |
| Metabolomic profile |  |  |  |  |  |  |  |  |
| Neither |  |  |  | Reference |  |  |  |  |
| Childhood | 0.024 | -0.089 | 0.138 | 0.811 | 0.014 | -0.098 | 0.126 | 0.811 |
| Adulthood | 0.060 | -0.029 | 0.150 | 0.542 | 0.019 | -0.071 | 0.109 | 0.811 |
| Both | 0.116 | 0.024 | 0.209 | 0.084 | 0.052 | -0.041 | 0.146 | 0.542 |
| Frailty index |  |  |  |  |  |  |  |  |
| Neither |  |  |  | Reference |  |  |  |  |
| Childhood | 0.224 | 0.204 | 0.244 | <0.001 | 0.221 | 0.201 | 0.241 | <0.001 |
| Adulthood | 0.207 | 0.191 | 0.222 | <0.001 | 0.168 | 0.152 | 0.183 | <0.001 |
| Both | 0.523 | 0.508 | 0.538 | <0.001 | 0.471 | 0.456 | 0.487 | <0.001 |
| Telomere length |  |  |  |  |  |  |  |  |
| Neither |  |  |  | Reference |  |  |  |  |
| Childhood | 0.015 | -0.008 | 0.038 | 0.315 | 0.018 | -0.005 | 0.042 | 0.248 |
| Adulthood | 0.005 | -0.013 | 0.023 | 0.673 | 0.003 | -0.015 | 0.021 | 0.731 |
| Both | 0.019 | 0.002 | 0.037 | 0.098 | 0.024 | 0.005 | 0.042 | 0.063 |
| Grip strength |  |  |  |  |  |  |  |  |
| Neither |  |  |  | Reference |  |  |  |  |
| Childhood | 0.011 | -0.001 | 0.024 | 0.093 | 0.010 | -0.003 | 0.022 | 0.123 |
| Adulthood | 0.040 | 0.031 | 0.050 | <0.001 | 0.026 | 0.016 | 0.035 | <0.001 |
| Both | 0.058 | 0.048 | 0.067 | <0.001 | 0.039 | 0.029 | 0.049 | <0.001 |
| Males |  |  |  |  |  |  |  |  |
| MileAge delta |  |  |  |  |  |  |  |  |
| Neither |  |  |  | Reference |  |  |  |  |
| Childhood | -0.013 | -0.045 | 0.020 | 0.708 | -0.012 | -0.045 | 0.021 | 0.708 |
| Adulthood | 0.004 | -0.025 | 0.033 | 0.943 | 0.001 | -0.029 | 0.031 | 0.946 |
| Both | 0.019 | -0.011 | 0.048 | 0.708 | 0.013 | -0.017 | 0.043 | 0.708 |
| Metabolomic profile |  |  |  |  |  |  |  |  |
| Neither |  |  |  | Reference |  |  |  |  |
| Childhood | 0.007 | -0.105 | 0.119 | 0.901 | -0.008 | -0.119 | 0.103 | 0.901 |
| Adulthood | 0.091 | -0.014 | 0.196 | 0.180 | 0.040 | -0.066 | 0.146 | 0.688 |
| Both | 0.163 | 0.058 | 0.269 | 0.014 | 0.125 | 0.019 | 0.232 | 0.063 |
| Frailty index |  |  |  |  |  |  |  |  |
| Neither |  |  |  | Reference |  |  |  |  |
| Childhood | 0.201 | 0.182 | 0.221 | <0.001 | 0.189 | 0.170 | 0.208 | <0.001 |
| Adulthood | 0.192 | 0.174 | 0.210 | <0.001 | 0.141 | 0.123 | 0.158 | <0.001 |
| Both | 0.456 | 0.439 | 0.474 | <0.001 | 0.384 | 0.366 | 0.401 | <0.001 |
| Telomere length |  |  |  |  |  |  |  |  |
| Neither |  |  |  | Reference |  |  |  |  |
| Childhood | 0.006 | -0.017 | 0.028 | 0.658 | 0.005 | -0.018 | 0.028 | 0.658 |
| Adulthood | -0.015 | -0.035 | 0.005 | 0.422 | -0.019 | -0.039 | 0.002 | 0.422 |
| Both | -0.006 | -0.026 | 0.014 | 0.658 | -0.009 | -0.029 | 0.012 | 0.658 |
| Grip strength |  |  |  |  |  |  |  |  |
| Neither |  |  |  | Reference |  |  |  |  |
| Childhood | -0.010 | -0.028 | 0.007 | 0.242 | -0.016 | -0.034 | 0.001 | 0.076 |
| Adulthood | 0.090 | 0.074 | 0.105 | <0.001 | 0.061 | 0.046 | 0.077 | <0.001 |
| Both | 0.089 | 0.073 | 0.104 | <0.001 | 0.046 | 0.030 | 0.061 | <0.001 |

*Note:* CI = confidence interval. Model 1–adjusted for chronological age; Model 2–adjusted for chronological age, ethnicity, highest educational/professional qualification, annual gross household income and neighbourhood deprivation. *P*-values shown are corrected for multiple testing using the Benjamini–Hochberg procedure (across exposure levels, ageing markers and models, separately for each sex). Sample sizes reported in Table S7.

### Childhood adversity item-specific associations

All types of childhood abuse (physical, emotional and sexual) were associated with a metabolite-predicted age exceeding chronological age in females (e.g., sexual abuse: *β* = 0.056, 95% CI 0.024-0.088, *p* = 0.002), whereas none of the corresponding estimates reached statistical significance in males (*p* > 0.126) (Figure 2; Table S8). However, no statistically significant sex-by-adversity interactions were detected (*p* _interaction_ > 0.186) (Table S9).

**Figure 2.**
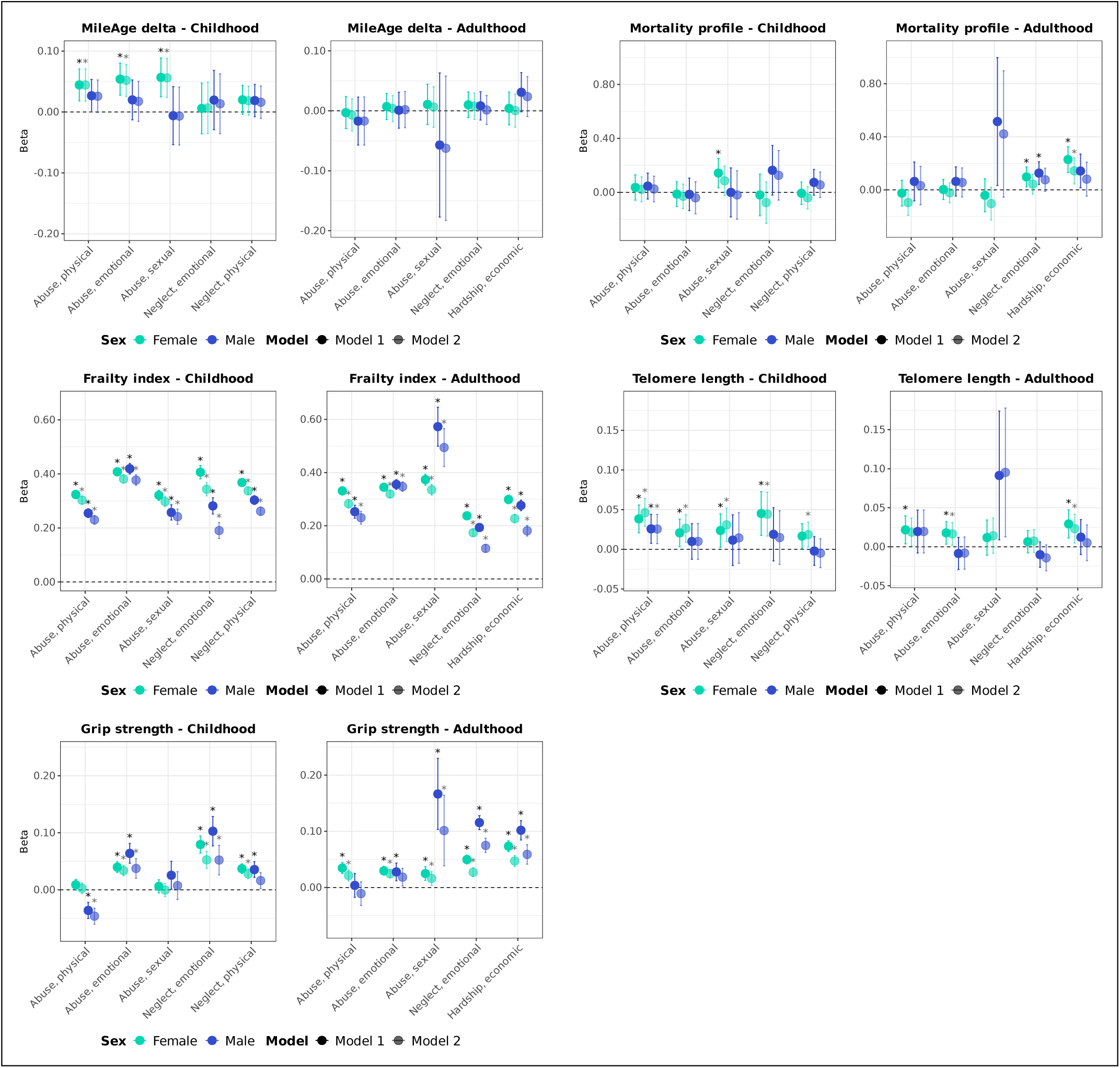
Ageing markers and adversity (item-specific). Sex-stratified associations between specific adversity items (adverse/traumatic events in childhood and in adulthood) and ageing markers (MileAge delta, frailty index, telomere length [reverse coded] and grip strength [reverse coded]). Estimates shown are ordinary least squares regression beta coefficients and 95% confidence intervals. Model 1–adjusted for chronological age; Model 2–adjusted for chronological age, ethnicity, highest educational/professional qualification, annual gross household income and neighbourhood deprivation. Asterisks indicate statistically significant associations, after correcting *p*-values for multiple testing using the Benjamini–Hochberg procedure (across childhood and adulthood exposures, ageing markers and models, separately for each sex). Sample sizes reported in Tables S2 and S5.

Childhood sexual abuse was associated with higher metabolomic mortality profile scores in age-adjusted models in females only (*β* = 0.143, 95% CI 0.034-0.252, *p* = 0.019) but the association attenuated after full adjustment (*β* = 0.085, 95% CI −0.023 to 0.194, *p* = 0.184). The interaction test was also not statistically significant (*p* _interaction_ = 0.481). Childhood adverse events were associated with greater frailty, with stronger associations observed in females (range: *β* = 0.297-0.381) than in males (range: *β* = 0.191-0.377), supported by statistically significant interaction tests (*p* _interaction_ ≤ 0.008). The exception was emotional abuse, for which there was no evidence of sex differences (females: *β* = 0.381, 95% CI 0.366-0.396, *p* < 0.001; males: *β* = 0.377, 95% CI 0.358-0.397, *p* < 0.001; *p* _interaction_ = 0.939).

Childhood physical abuse was associated with shorter telomeres in both sexes (females: *β* = 0.046, 95% CI 0.029-0.064, *p* < 0.001; males: *β* = 0.026, 95% CI 0.007-0.044, *p* = 0.017). All other childhood adverse events were associated with shorter telomeres only in females (range: *β* = 0.019-0.044), though none of the interaction tests reached statistical significance (*p* _interaction_ ζ 0.084). Childhood physical abuse was associated with greater grip strength in males only (*β* = −0.046, 95% CI −0.060 to −0.032, *p* < 0.001), yielding a statistically significant interaction (*β* _interaction_ = −0.05, 95% CI −0.06 to −0.03, *p* < 0.001). Physical neglect was associated with weaker grip strength in females but in males the association did not reach statistical significance after full adjustment (*β* = 0.016, 95% CI 0.002-0.030, *p* = 0.055). No other interaction tests were statistically significant.

### Adulthood adversity item-specific associations

Adulthood economic hardship was associated with higher metabolomic mortality profile scores in females (*β* = 0.144, 95% CI 0.045-0.242, *p* = 0.009) but not in males (*β* = 0.081, 95% CI −0.047 to 0.209, *p* = 0.368) (Figure 2; Table S10). However, the sex-by-adversity interaction test was not statistically significant (*p* _interaction_ = 0.534) (Table S11). We observed stronger associations with frailty for adulthood physical abuse, emotional neglect and economic hardship in females (*p* _interaction_ ≤ 0.021), whereas sexual and emotional abuse were more strongly associated with frailty in males (*p* _interaction_ ≤ 0.023). Adulthood emotional abuse (*β* = 0.017, 95% CI 0.002-0.031, *p* = 0.043) and economic hardship (*β* = 0.023, 95% CI 0.005-0.042, *p* = 0.025) were associated with shorter telomeres in females only, with an interaction detected for emotional abuse (*β* _interaction_ = −0.03, 95% CI −0.06 to −0.01, *p* = 0.034). Adulthood physical abuse was associated with weaker grip strength in females only (*β* = 0.021, 95% CI 0.012-0.031, *p* < 0.001) but the interaction test was not statistically significant after full adjustment (*β* _interaction_ = −0.02, 95% CI −0.04 to 0.00, *p* = 0.141). Associations of adulthood sexual abuse, emotional neglect and economic hardship with grip strength were stronger in males. All interaction tests were statistically significant (*p* _interaction_ < 0.001), except for economic hardship (*β* _interaction_ = 0.02, 95% CI 0.00-0.04, *p* = 0.116).

## Discussion

In this large population-based study, we found that sex differences in the associations between adversity and markers of biological ageing vary by timing of adversity exposure. Childhood adversity was more strongly associated with older biological ageing profiles in females than in males, including advanced metabolomic ageing, greater frailty, shorter telomeres and weaker grip strength. In contrast, adulthood adversity showed stronger associations with certain ageing markers in males, notably greater frailty and weaker grip strength. This pattern suggests that males and females may have different sensitive periods of vulnerability to adversity, with the timing and type of adversity each shaping distinct patterns of association by sex. For example, childhood sexual abuse in females was associated with older biological ageing profiles across most markers, while adulthood sexual abuse in males was more strongly associated with weaker grip strength. These findings underscore the importance of considering both timing and type of adversity in studying the links between adversity and biological ageing.

Our findings support a growing body of research showing that both childhood and adulthood adversity are associated with physical and mental health outcomes and markers linked to biological ageing. For example, prior studies have identified associations between cumulative trauma exposure and the development of psychopathology (Hakamata, Suzuki, Kobashikawa, & Hori, 2022; Ogle, Rubin, & Siegler, 2014) and associations with biological ageing markers such as telomere shortening (Ridout et al., 2018), reduced grip strength (Duchowny, Hicken, Cawthon, Glymour, & Clarke, 2020), greater frailty (Yang et al., 2024) and advanced metabolomic ageing (Aas et al., 2025). However, few investigations have examined sex-specific patterns in associations between adversity and biological ageing, despite evidence suggesting that males and females differ both in their level of exposure to adversity and possibly in their biological vulnerability to its long-term effects (Garvin & Bolton, 2022).

Our study addresses this gap by showing that the timing of adversity alters the direction of sex differences across certain ageing markers, supported by a combination of sex-stratified analyses and formal sex-by-adversity interaction testing.

Sex hormones play a crucial role in shaping biological resilience and vulnerability, with oestrogen providing significant protective effects in females (Huang, Li, Liang, Chen, & Tang, 2024). Although females often report greater exposure to adversity, oestrogen may modulate stress responses, reduce inflammation and support neuroplasticity (Galea et al., 2017). Oestrogen also may activate telomerase and has antioxidant properties that help protect telomeres from oxidative stress (Viña, Borrás, Gambini, Sastre, & Pallardó, 2005). This partly explains why females generally have longer telomeres. Testosterone, which is more abundant in males, lacks antioxidant properties and may indeed exacerbate oxidative stress on telomeres (Alonso-Alvarez, Bertrand, Faivre, Chastel, & Sorci, 2007; Axson et al., 2018). Oestrogen may also help preserve muscle and bone health, thereby contributing to resilience against frailty (Gordon & Hubbard, 2018). In relation to muscle physiology, testosterone also plays a critical role. Higher testosterone levels are strongly associated with increased muscle mass, which directly contributes to greater grip strength (Chiu, Shih, & Chen, 2020). Adequate levels of testosterone help maintain muscular function, and reductions can lead to muscle loss and diminished strength (Le Noan-Lainé et al., 2024).

Nonetheless, the observation that females showed stronger associations for childhood adversity across several ageing markers suggests that hormonal explanations alone are insufficient. The Childhood Trauma Screener items used here capture predominantly relational and family-based adversity, such as emotional and sexual abuse. Females are more likely to report some of these experiences, particularly sexual abuse, which in our study was consistently associated with older biological ageing profiles. Greater exposure to, and possibly differential reporting of, specific types of adversity could partly explain the stronger associations in females. In contrast, the stronger adulthood adversity associations in males may reflect that some adulthood adverse events, such as partner violence, are less frequently reported by males, meaning that those who do report them could represent a subgroup with particularly severe exposures.

Our study is, to our knowledge, the first to investigate potential sex differences in both the timing (whether in childhood or adulthood) and type of adversity exposure in relation to multiple biological ageing markers. While several differences were supported by statistically significant sex-by-adversity interaction tests, others were based on patterns in stratified analyses and should be interpreted cautiously. Future research should investigate whether these associations are further moderated by hormonal status, including whether adulthood adversity occurred before or after menopause in females, which we could not investigate here. Incorporating levels of sex hormones (testosterone and oestrogen) could help clarify the biological mechanisms underlying observed sex differences. Our study primarily focused on relational forms of adversity, except for one question about financial hardship in adulthood. Studying non-relational traumatic experiences, such as being exposed to war or a natural disaster, remains important to test whether our findings generalise or whether they are specific to the types of adverse events investigated here (Hoppen, Morina, Aas, & Mutz, 2025). Additionally, future research should examine the extent to which cognitive and emotional coping mechanisms, as well as the presence of mental disorders (Mutz, Gilchrist, Allegrini, Sanchez Roige, & Lewis, 2025; Mutz, Hoppen, Fabbri, & Lewis, 2022), mediate the relationship between adversity and biological ageing, and how this relationship is differentially expressed in females and males.

A strength of this study is the large sample size, with over 150,000 individuals, which provided substantial statistical power to detect even modest sex-specific associations and sex-by-adversity interactions. The inclusion of a wide array of biological ageing markers expands previous findings that often focused on a single ageing biomarker. However, the large sample also means that some statistically significant findings may represent small absolute differences, and their clinical relevance should be interpreted cautiously.

Several limitations should be noted. First, the measurement of telomere length, widely used as a biomarker of cellular aging, has inherent constraints. Specifically, our analysis is based on mean telomere length, which does not capture the distribution of telomere lengths or the presence and proportion of critically short telomeres, which are thought to have greater biological relevance for ageing. Second, we relied on a short version of the Childhood Trauma Questionnaire to capture relational adversity in childhood, omitting other types of adverse events, such as peer bullying, parental divorce or separation and exposure to community violence. Third, participants were asked to recall adverse events retrospectively, which introduces the possibility of recall bias and may result in underreporting or misclassification. However, retrospective, even more so than prospective, collection of adverse events, has been identified as a marker of poorer functioning in adulthood (Danese & Widom, 2023), indicating that perception matters greatly in this context. A recent meta-analysis suggested low overlap between prospective and retrospective measures, with the latter indeed having higher sensitivity in detecting cases that otherwise could go unnoticed (Baldwin, Reuben, Newbury, & Danese, 2019). It is also possible that recall accuracy or reporting tendencies differ by sex, which could influence observed patterns and should be addressed in future studies.

### Conclusion

This study identified sex-specific patterns in the associations between adversity and biological ageing markers in a large, population-based sample. The timing of adversity was critical: childhood adversity was more strongly linked to ageing markers in females, whereas adulthood adversity showed stronger associations in males. Several of these differences were supported by statistically significant sex-by-adversity interactions, while others were observed only in stratified analyses and require replication. Our findings suggest that sex differences in ageing profiles linked to adversity may emerge from distinct sensitive periods of vulnerability, highlighting the need to integrate both sex and timing of exposure into studies of adversity and biological ageing.

## Data Availability

The data used are available to all bona fide researchers for health-related research that is in the public interest, subject to an application process and approval criteria. Study materials are publicly available online at http://www.ukbiobank.ac.uk.

## Acknowledgements

JM is funded by the King’s Prize Fellowship. MA is funded by the MRC fellowship (#MR/W027720/1). Computational analyses were supported by King’s Computational Research, Engineering and Technology Environment (CREATE). This research has been conducted using data from UK Biobank. Data access permission has been granted under UK Biobank application 45514.

## Financial disclosures

The authors declare no conflicts of interest.

## Authorship contributions

JM and MA conceived the idea of the study. JM acquired the data and performed the statistical analysis. JM, LDB and MA wrote the manuscript. JM, LDB, THH, NM and MA interpreted the findings and revised the manuscript. All authors read and approved the final manuscript.

## Ethics

Ethical approval for the UK Biobank study has been granted by the National Information Governance Board for Health and Social Care and the NHS North West Multicentre Research Ethics Committee (11/NW/0382). No project-specific ethical approval is needed.

## Supplementary material

Supplementary information is available online.

## Notes

### Competing Interest Statement

The authors have declared no competing interest.

