## Supplementary material for "Sex differences in associations between adversity and biological ageing"

This material accompanies the article

### 1. Sample characteristics childhood adversity

**Table S1.** Sample characteristics stratified by childhood adversity and sex

| | Childhood adversity | | | | |
|---|---|---|---|---|---|
| | Female | | Male | | |
| | No (*N*=50,894) | Yes (*N*=35,578) | No (*N*=39,597) | Yes (*N*=27,488) | Full sample (*N*=153,557) |
| MileAge delta, mean (SD)[1] | 0.12 (3.77) | 0.22 (3.84) | -0.29 (3.69) | -0.25 (3.73) | -0.03 (3.76) |
| Mortality profile, mean (SD)[1] | -45.17 (0.51) | -45.16 (0.53) | -45.03 (0.53) | -45.02 (0.51) | -45.11 (0.52) |
| Frailty index, mean (SD)[1] | 0.10 (0.06) | 0.13 (0.07) | 0.10 (0.06) | 0.12 (0.07) | 0.11 (0.07) |
| T/S ratio, mean (SD)[1] | 0.14 (0.98) | 0.14 (0.99) | -0.07 (0.99) | -0.05 (0.99) | 0.05 (0.99) |
| Grip strength, mean (SD)[1] | 25.41 (6.35) | 25.46 (6.62) | 41.45 (8.79) | 41.57 (9.02) | 32.45 (11.03) |
| Age, mean (SD) | 56.43 (7.59) | 55.19 (7.66) | 57.42 (7.72) | 56.50 (7.85) | 56.41 (7.73) |
| Ethnicity | | | | | |
| White | 49,835 (97.9%) | 33,959 (95.4%) | 38,714 (97.8%) | 26,230 (95.4%) | 148,738 (96.9%) |
| Mixed | 190 (0.4%) | 318 (0.9%) | 102 (0.3%) | 182 (0.7%) | 792 (0.5%) |
| Black | 205 (0.4%) | 421 (1.2%) | 152 (0.4%) | 289 (1.1%) | 1,067 (0.7%) |
| Asian | 262 (0.5%) | 299 (0.8%) | 308 (0.8%) | 392 (1.4%) | 1,261 (0.8%) |
| Chinese | 89 (0.2%) | 160 (0.4%) | 29 (0.1%) | 71 (0.3%) | 349 (0.2%) |
| Other | 208 (0.4%) | 303 (0.9%) | 136 (0.3%) | 190 (0.7%) | 837 (0.5%) |
| Missing[2] | 105 (0.2%) | 118 (0.3%) | 156 (0.4%) | 134 (0.5%) | 513 (0.3%) |
| Highest qualification | | | | | |
| None | 3,371 (6.6%) | 2,330 (6.5%) | 2,551 (6.4%) | 2,192 (8.0%) | 10,444 (6.8%) |
| O levels/GCSEs/CSEs | 13,109 (25.8%) | 8,767 (24.6%) | 8,004 (20.2%) | 6,015 (21.9%) | 35,895 (23.4%) |
| A levels/NVQ/HND/HNC[3] | 11,804 (23.2%) | 8,255 (23.2%) | 9,243 (23.3%) | 6,775 (24.6%) | 36,077 (23.5%) |
| Degree | 22,130 (43.5%) | 15,870 (44.6%) | 19,458 (49.1%) | 12,250 (44.6%) | 69,708 (45.4%) |
| Missing[2] | 480 (0.9%) | 356 (1.0%) | 341 (0.9%) | 256 (0.9%) | 1,433 (0.9%) |
| Household income[4] | | | | | |
| Very low | 6,291 (12.4%) | 5,413 (15.2%) | 3,765 (9.5%) | 3,356 (12.2%) | 18,825 (12.3%) |
| Low | 10,967 (21.5%) | 7,726 (21.7%) | 8,004 (20.2%) | 5,564 (20.2%) | 32,261 (21.0%) |
| Medium | 12,678 (24.9%) | 8,987 (25.3%) | 10,841 (27.4%) | 7,610 (27.7%) | 40,116 (26.1%) |
| High | 11,025 (21.7%) | 7,430 (20.9%) | 10,793 (27.3%) | 6,950 (25.3%) | 36,198 (23.6%) |
| Very high | 3,320 (6.5%) | 2,157 (6.1%) | 3,484 (8.8%) | 2,145 (7.8%) | 11,106 (7.2%) |
| Missing[2] | 6,613 (13.0%) | 3,865 (10.9%) | 2,710 (6.8%) | 1,863 (6.8%) | 15,051 (9.8%) |
| Neighbourhood deprivation | | | | | |
| Q1 | 11,993 (23.6%) | 7,060 (19.8%) | 9,849 (24.9%) | 5,901 (21.5%) | 34,803 (22.7%) |
| Q2 | 11,395 (22.4%) | 6,980 (19.6%) | 9,056 (22.9%) | 5,619 (20.4%) | 33,050 (21.5%) |
| Q3 | 10,786 (21.2%) | 7,163 (20.1%) | 8,179 (20.7%) | 5,510 (20.0%) | 31,638 (20.6%) |
| Q4 | 9,780 (19.2%) | 7,544 (21.2%) | 7,184 (18.1%) | 5,575 (20.3%) | 30,083 (19.6%) |
| Q5 | 6,884 (13.5%) | 6,781 (19.1%) | 5,278 (13.3%) | 4,845 (17.6%) | 23,788 (15.5%) |
| Missing[2] | 56 (0.1%) | 50 (0.1%) | 51 (0.1%) | 38 (0.1%) | 195 (0.1%) |

*Note:* Numbers shown are counts and percentages unless indicated otherwise. SD = standard deviation. GCSEs = general certificate of secondary education; CSE = certificate of secondary education; NVQ = national vocational qualification; HND = higher national diploma; HNC = higher national certificate. [1] Sample sizes for health and ageing markers: $n$ = 69,451 (MileAge delta), 4,260 (mortality profile), 153,387 (frailty index), 144,968 (T/S ratio) and 153,055 (grip strength). [2] Missing data may also include “do not know” or “prefer not to answer”. [3] Also includes 'other professional qualifications'. [4] Annual household income groups: very low (<£18,000), low (£18,000–£30,999), middle (£31,000–£51,999), high (£52,000–£100,000) and very high (>£100,000).

#### 2. Sample sizes childhood adversity

**Table S2.** Childhood adversity analytical sample sizes

| Childhood adversity | MileAge delta | Mortality profile | Frailty index | Telomere length | Grip strength |
|---|---|---|---|---|---|
| Females | | | | | |
| Sum score[1] | 0.73 (1.10) | 0.68 (1.05) | 0.74 (1.11) | 0.74 (1.11) | 0.74 (1.11) |
| Specific items | | | | | |
| Abuse, physical | 32,592 (82.7%) | 2,091 (84.4%) | 73,356 (82.7%) | 69,116 (82.7%) | 73,193 (82.7%) |
| Yes | 6,829 (17.3%) | 387 (15.6%) | 15,353 (17.3%) | 14,469 (17.3%) | 15,321 (17.3%) |
| Abuse, emotional | 32,514 (82.5%) | 2,080 (83.9%) | 72,834 (82.2%) | 68,650 (82.2%) | 72,670 (82.2%) |
| Yes | 6,889 (17.5%) | 399 (16.1%) | 15,804 (17.8%) | 14,863 (17.8%) | 15,771 (17.8%) |
| Abuse, sexual | 34,602 (89.0%) | 2,177 (89.1%) | 77,835 (88.9%) | 73,309 (88.9%) | 77,669 (89.0%) |
| Yes | 4,271 (11.0%) | 265 (10.9%) | 9,674 (11.1%) | 9,139 (11.1%) | 9,646 (11.0%) |
| Neglect, emotional | 36,893 (94.0%) | 2,347 (95.0%) | 82,840 (93.9%) | 78,048 (93.9%) | 82,659 (93.9%) |
| Yes | 2,364 (6.0%) | 124 (5.0%) | 5,418 (6.1%) | 5,106 (6.1%) | 5,400 (6.1%) |
| Neglect, physical | 30,389 (77.2%) | 1,949 (78.9%) | 68,089 (76.9%) | 64,190 (76.9%) | 67,944 (76.9%) |
| Yes | 8,999 (22.8%) | 522 (21.1%) | 20,503 (23.1%) | 19,297 (23.1%) | 20,453 (23.1%) |
| Males | | | | | |
| Sum score[1] | 0.65 (0.96) | 0.60 (0.90) | 0.66 (0.97) | 0.66 (0.97) | 0.66 (0.97) |
| Specific items | | | | | |
| Abuse, physical | 24,808 (78.9%) | 1,498 (79.9%) | 53,644 (78.9%) | 50,879 (78.9%) | 53,542 (78.9%) |
| Yes | 6,642 (21.1%) | 376 (20.1%) | 14,377 (21.1%) | 13,633 (21.1%) | 14,342 (21.1%) |
| Abuse, emotional | 27,455 (87.3%) | 1,654 (88.3%) | 59,328 (87.2%) | 56,256 (87.2%) | 59,205 (87.2%) |
| Yes | 3,984 (12.7%) | 220 (11.7%) | 8,671 (12.8%) | 8,239 (12.8%) | 8,657 (12.8%) |
| Abuse, sexual | 29,593 (94.5%) | 1,775 (95.3%) | 63,793 (94.2%) | 60,511 (94.2%) | 63,660 (94.2%) |
| Yes | 1,727 (5.5%) | 88 (4.7%) | 3,939 (5.8%) | 3,735 (5.8%) | 3,934 (5.8%) |
| Neglect, emotional | 29,688 (94.7%) | 1,781 (95.4%) | 64,205 (94.8%) | 60,894 (94.7%) | 64,072 (94.7%) |
| Yes | 1,655 (5.3%) | 86 (4.6%) | 3,553 (5.2%) | 3,381 (5.3%) | 3,552 (5.3%) |
| Neglect, physical | 24,684 (78.7%) | 1,490 (79.5%) | 53,169 (78.3%) | 50,433 (78.3%) | 53,053 (78.3%) |
| Yes | 6,690 (21.3%) | 384 (20.5%) | 14,700 (21.7%) | 13,939 (21.7%) | 14,679 (21.7%) |

*Note:* Numbers shown are counts and percentages unless indicated otherwise. [1] mean and standard deviation.

#### 3. Sex-by-adversity interactions

**Table S3.** Sex interactions for associations between adverse events and ageing markers

| | Model 1 (adj. age) | | | | Model 2 (full adjustment) | | | |
|---|---|---|---|---|---|---|---|---|
| Childhood | *β* | 95% CI | | *p* | *β* | 95% CI | | *p* |
| MileAge delta | 0.00 | -0.01 | 0.02 | 0.737 | 0.00 | -0.01 | 0.02 | 0.739 |
| Metabolomic profile | 0.01 | -0.04 | 0.07 | 0.737 | 0.02 | -0.04 | 0.07 | 0.717 |
| Frailty index | 0.00 | -0.01 | 0.00 | 0.499 | -0.01 | -0.02 | 0.00 | 0.063 |
| Telomere length | -0.01 | -0.02 | 0.00 | 0.272 | -0.01 | -0.02 | 0.00 | 0.133 |
| Grip strength | 0.00 | -0.01 | 0.01 | 0.737 | 0.00 | -0.01 | 0.00 | 0.276 |
| Adulthood | | | | | | | | |
| MileAge delta | 0.01 | 0.00 | 0.03 | 0.272 | 0.01 | -0.01 | 0.03 | 0.276 |
| Metabolomic profile | 0.04 | -0.02 | 0.10 | 0.272 | 0.04 | -0.01 | 0.10 | 0.272 |
| Frailty index | 0.02 | 0.01 | 0.03 | <0.001 | 0.01 | 0.00 | 0.02 | 0.017 |
| Telomere length | -0.01 | -0.02 | 0.00 | 0.096 | -0.01 | -0.02 | 0.00 | 0.063 |
| Grip strength | 0.03 | 0.03 | 0.04 | <0.001 | 0.03 | 0.02 | 0.03 | <0.001 |

*Note:* CI = confidence interval. Model 1–adjusted for chronological age; Model 2–adjusted for chronological age, ethnicity, highest educational/professional qualification, annual gross household income and neighbourhood deprivation. *P*-values shown are corrected for multiple testing using the Benjamini–Hochberg procedure (across childhood and adulthood exposures, ageing markers and models). Sample sizes reported in Tables S1 and S4.

### 4. Sample characteristics adulthood adversity

**Table S4.** Sample characteristics stratified by adulthood adversity and sex

| | Adulthood adversity | | | | |
|---|---|---|---|---|---|
| | Female | | Male | | |
| | No (*N*=35,764) | Yes (*N*=49,065) | No (*N*=34,189) | Yes (*N*=31,830) | Full sample (*N*=150,848) |
| MileAge delta, mean (SD)[1] | 0.14 (3.76) | 0.18 (3.82) | -0.31 (3.71) | -0.23 (3.70) | -0.03 (3.76) |
| Mortality profile, mean (SD)[1] | -45.19 (0.49) | -45.14 (0.54) | -45.05 (0.52) | -45.00 (0.53) | -45.10 (0.52) |
| Frailty index, mean (SD)[1] | 0.10 (0.06) | 0.12 (0.07) | 0.10 (0.06) | 0.12 (0.07) | 0.11 (0.07) |
| T/S ratio, mean (SD)[1] | 0.14 (0.99) | 0.14 (0.98) | -0.07 (0.98) | -0.04 (0.99) | 0.05 (0.99) |
| Grip strength, mean (SD)[1] | 25.67 (6.34) | 25.29 (6.55) | 41.93 (8.72) | 41.10 (9.04) | 32.49 (11.03) |
| Age, mean (SD) | 56.19 (7.55) | 55.66 (7.68) | 57.42 (7.58) | 56.56 (7.96) | 56.38 (7.72) |
| Ethnicity | | | | | |
| White | 35,131 (98.2%) | 47,084 (96.0%) | 33,515 (98.0%) | 30,411 (95.5%) | 146,141 (96.9%) |
| Mixed | 149 (0.4%) | 363 (0.7%) | 120 (0.4%) | 157 (0.5%) | 789 (0.5%) |
| Black | 88 (0.2%) | 533 (1.1%) | 108 (0.3%) | 324 (1.0%) | 1,053 (0.7%) |
| Asian | 137 (0.4%) | 396 (0.8%) | 203 (0.6%) | 480 (1.5%) | 1,216 (0.8%) |
| Chinese | 56 (0.2%) | 183 (0.4%) | 30 (0.1%) | 71 (0.2%) | 340 (0.2%) |
| Other | 129 (0.4%) | 364 (0.7%) | 92 (0.3%) | 225 (0.7%) | 810 (0.5%) |
| Missing[2] | 74 (0.2%) | 142 (0.3%) | 121 (0.4%) | 162 (0.5%) | 499 (0.3%) |
| Highest qualification | | | | | |
| None | 2,059 (5.8%) | 3,458 (7.0%) | 1,871 (5.5%) | 2,723 (8.6%) | 10,111 (6.7%) |
| O levels/GCSEs/CSEs | 8,559 (23.9%) | 12,828 (26.1%) | 6,799 (19.9%) | 6,988 (22.0%) | 35,174 (23.3%) |
| A levels/NVQ/HND/HNC[3] | 8,216 (23.0%) | 11,454 (23.3%) | 8,016 (23.4%) | 7,715 (24.2%) | 35,401 (23.5%) |
| Degree | 16,643 (46.5%) | 20,805 (42.4%) | 17,228 (50.4%) | 14,097 (44.3%) | 68,773 (45.6%) |
| Missing[2] | 287 (0.8%) | 520 (1.1%) | 275 (0.8%) | 307 (1.0%) | 1,389 (0.9%) |
| Household income[4] | | | | | |
| Very low | 3,258 (9.1%) | 8,192 (16.7%) | 2467 (7.2%) | 4,445 (14.0%) | 18,362 (12.2%) |
| Low | 7,179 (20.1%) | 11,173 (22.8%) | 6,335 (18.5%) | 6,933 (21.8%) | 31,620 (21.0%) |
| Medium | 9,156 (25.6%) | 12,178 (24.8%) | 9,452 (27.6%) | 8,775 (27.6%) | 39,561 (26.2%) |
| High | 8,944 (25.0%) | 9,284 (18.9%) | 10,295 (30.1%) | 7,298 (22.9%) | 35,821 (23.7%) |
| Very high | 2,902 (8.1%) | 2,511 (5.1%) | 3,496 (10.2%) | 2,092 (6.6%) | 11,001 (7.3%) |
| Missing[2] | 4,325 (12.1%) | 5,727 (11.7%) | 2,144 (6.3%) | 2,287 (7.2%) | 14,483 (9.6%) |
| Neighbourhood deprivation | | | | | |
| Q1 | 8,994 (25.1%) | 9,668 (19.7%) | 8,959 (26.2%) | 6,517 (20.5%) | 34,138 (22.6%) |
| Q2 | 8,310 (23.2%) | 9,695 (19.8%) | 8,160 (23.9%) | 6,283 (19.7%) | 32,448 (21.5%) |
| Q3 | 7,639 (21.4%) | 9,990 (20.4%) | 7,080 (20.7%) | 6,367 (20.0%) | 31,076 (20.6%) |
| Q4 | 6,546 (18.3%) | 10,458 (21.3%) | 6,103 (17.9%) | 6,465 (20.3%) | 29,572 (19.6%) |
| Q5 | 4,235 (11.8%) | 9,190 (18.7%) | 3,850 (11.3%) | 6,147 (19.3%) | 23,422 (15.5%) |
| Missing[2] | 40 (0.1%) | 64 (0.1%) | 37 (0.1%) | 51 (0.2%) | 192 (0.1%) |

*Note:* Numbers shown are counts and percentages unless indicated otherwise. SD = standard deviation. GCSEs = general certificate of secondary education; CSE = certificate of secondary education; NVQ = national vocational qualification; HND = higher national diploma; HNC = higher national certificate. [1] Sample sizes for health and ageing markers: $n$ = 68,126 (MileAge delta), 4,205 (mortality profile), 150,690 (frailty index), 142,381 (T/S ratio) and 150,362 (grip strength). [2] Missing data may also include “do not know” or “prefer not to answer”. [3] Also includes 'other professional qualifications'. [4] Annual household income groups: very low (<£18,000), low (£18,000–£30,999), middle (£31,000–£51,999), high (£52,000–£100,000) and very high (>£100,000).

### 5. Sample sizes adulthood adversity

**Table S5.** Adulthood adversity analytical sample sizes

| Adulthood adversity | MileAge delta | Mortality profile | Frailty index | Telomere length | Grip strength |
|---|---|---|---|---|---|
| Females | | | | | |
| Sum score[1] | 1.05 (1.20) | 0.93 (1.13) | 1.05 (1.19) | 1.05 (1.19) | 1.05 (1.19) |
| Specific items | | | | | |
| Abuse, physical | 32,816 (83.4%) | 2,123 (85.8%) | 74,039 (83.6%) | 69,710 (83.5%) | 73,865 (83.6%) |
| Yes | 6,542 (16.6%) | 352 (14.2%) | 14,546 (16.4%) | 13,765 (16.5%) | 14,522 (16.4%) |
| Abuse, emotional | 27,393 (69.6%) | 1,795 (72.5%) | 61,779 (69.7%) | 58,153 (69.6%) | 61,631 (69.7%) |
| Yes | 11,980 (30.4%) | 681 (27.5%) | 26,839 (30.3%) | 25,341 (30.4%) | 26,788 (30.3%) |
| Abuse, sexual | 35,549 (90.3%) | 2,276 (92.0%) | 80,060 (90.4%) | 75,439 (90.4%) | 79,878 (90.4%) |
| Yes | 3,799 (9.7%) | 198 (8.0%) | 8,475 (9.6%) | 7,986 (9.6%) | 8,463 (9.6%) |
| Neglect, emotional | 25,857 (67.5%) | 1,690 (69.9%) | 58,129 (67.4%) | 54,828 (67.5%) | 57,976 (67.4%) |
| Yes | 12,447 (32.5%) | 727 (30.1%) | 28,056 (32.6%) | 26,379 (32.5%) | 28,011 (32.6%) |
| Economic hardship | 32,684 (84.0%) | 2,111 (86.2%) | 73,512 (84.0%) | 69,280 (84.0%) | 73,341 (84.0%) |
| Yes | 6,214 (16.0%) | 338 (13.8%) | 13,990 (16.0%) | 13,157 (16.0%) | 13,963 (16.0%) |
| Males | | | | | |
| Sum score[1] | 0.68 (0.85) | 0.62 (0.84) | 0.69 (0.86) | 0.69 (0.86) | 0.69 (0.86) |
| Specific items | | | | | |
| Abuse, physical | 28,926 (92.0%) | 1,736 (92.5%) | 62,529 (91.9%) | 59,308 (91.9%) | 62,402 (91.9%) |
| Yes | 2,511 (8.0%) | 140 (7.5%) | 5,485 (8.1%) | 5,195 (8.1%) | 5,474 (8.1%) |
| Abuse, emotional | 26,530 (84.4%) | 1,604 (85.5%) | 57,287 (84.3%) | 54,323 (84.2%) | 57,164 (84.3%) |
| Yes | 4,900 (15.6%) | 272 (14.5%) | 10,702 (15.7%) | 10,157 (15.8%) | 10,686 (15.7%) |
| Abuse, sexual | 31,191 (99.2%) | 1,865 (99.4%) | 67,475 (99.2%) | 63,990 (99.2%) | 67,337 (99.2%) |
| Yes | 258 (0.8%) | 12 (0.6%) | 564 (0.8%) | 534 (0.8%) | 564 (0.8%) |
| Neglect, emotional | 21,146 (68.5%) | 1,304 (70.9%) | 45,812 (68.6%) | 43,433 (68.6%) | 45,716 (68.6%) |
| Yes | 9,708 (31.5%) | 536 (29.1%) | 20,982 (31.4%) | 19,904 (31.4%) | 20,942 (31.4%) |
| Economic hardship | 27,051 (87.2%) | 1,667 (89.8%) | 58,472 (87.1%) | 55,426 (87.0%) | 58,346 (87.1%) |
| Yes | 3,985 (12.8%) | 190 (10.2%) | 8,680 (12.9%) | 8,259 (13.0%) | 8,671 (12.9%) |

*Note:* Numbers shown are counts and percentages unless indicated otherwise. [1] mean and standard deviation.

#### 6. Sample sizes cross-classification

**Table S6.** Childhood and adulthood adversity analytical sample sizes

| Cross-classification | MileAge delta | Mortality profile | Frailty index | Telomere length | Grip strength |
|---|---|---|---|---|---|
| Overall | | | | | |
| Neither | 21,921 (32.7%) | 1,446 (35.1%) | 48,284 (32.6%) | 45,627 (32.6%) | 48,156 (32.6%) |
| Childhood | 9,464 (14.1%) | 638 (15.5%) | 20,745 (14.0%) | 19,618 (14.0%) | 20,697 (14.0%) |
| Adulthood | 17,691 (26.4%) | 1,053 (25.5%) | 38,843 (26.3%) | 36,736 (26.3%) | 38,762 (26.3%) |
| Both | 17,889 (26.7%) | 987 (23.9%) | 40,086 (27.1%) | 37,846 (27.1%) | 40,018 (27.1%) |
| Females | | | | | |
| Neither | 11,291 (30.6%) | 756 (32.5%) | 25,229 (30.5%) | 23,787 (30.5%) | 25,162 (30.4%) |
| Childhood | 4,399 (11.9%) | 313 (13.5%) | 9,940 (12.0%) | 9,348 (12.0%) | 9,913 (12.0%) |
| Adulthood | 10,495 (28.5%) | 660 (28.4%) | 23,448 (28.3%) | 22,106 (28.3%) | 23,397 (28.3%) |
| Both | 10,677 (29.0%) | 594 (25.6%) | 24,234 (29.3%) | 22,829 (29.2%) | 24,188 (29.3%) |
| Males | | | | | |
| Neither | 10,630 (35.3%) | 690 (38.3%) | 23,055 (35.4%) | 21,840 (35.4%) | 22,994 (35.4%) |
| Childhood | 5,065 (16.8%) | 325 (18.0%) | 10,805 (16.6%) | 10,270 (16.6%) | 10,784 (16.6%) |
| Adulthood | 7,196 (23.9%) | 393 (21.8%) | 15,395 (23.6%) | 14,630 (23.7%) | 15,365 (23.6%) |
| Both | 7,212 (24.0%) | 393 (21.8%) | 15,852 (24.3%) | 15,017 (24.3%) | 15,830 (24.4%) |

*Note:* Numbers shown are counts and percentages.

#### 7. Sex-by-adversity interactions cross-classification

**Table S7.** Sex interactions for associations between adverse events and ageing markers

| | Model 1 (adj. age) | | | | Model 2 (full adjustment) | | | |
|---|---|---|---|---|---|---|---|---|
| MileAge delta | *β* | 95% CI | | *p* | *β* | 95% CI | | *p* |
| Neither | Reference | | | | | | | |
| Childhood | -0.01 | -0.06 | 0.04 | 0.797 | -0.01 | -0.06 | 0.04 | 0.797 |
| Adulthood | 0.02 | -0.02 | 0.06 | 0.499 | 0.02 | -0.02 | 0.06 | 0.499 |
| Both | 0.00 | -0.04 | 0.04 | 0.855 | 0.00 | -0.04 | 0.04 | 0.894 |
| Metabolomic profile | | | | | | | | |
| Neither | Reference | | | | | | | |
| Childhood | -0.03 | -0.19 | 0.13 | 0.797 | -0.03 | -0.19 | 0.13 | 0.797 |
| Adulthood | 0.03 | -0.11 | 0.17 | 0.797 | 0.02 | -0.12 | 0.16 | 0.823 |
| Both | 0.03 | -0.11 | 0.17 | 0.797 | 0.05 | -0.09 | 0.19 | 0.723 |
| Frailty index | | | | | | | | |
| Neither | Reference | | | | | | | |
| Childhood | -0.02 | -0.05 | 0.01 | 0.257 | -0.03 | -0.06 | 0.00 | 0.084 |
| Adulthood | -0.01 | -0.04 | 0.01 | 0.426 | -0.02 | -0.04 | 0.00 | 0.155 |
| Both | -0.07 | -0.09 | -0.04 | <0.001 | -0.08 | -0.10 | -0.06 | <0.001 |
| Telomere length | | | | | | | | |
| Neither | Reference | | | | | | | |
| Childhood | -0.01 | -0.05 | 0.02 | 0.572 | -0.02 | -0.05 | 0.01 | 0.499 |
| Adulthood | -0.02 | -0.05 | 0.00 | 0.189 | -0.03 | -0.05 | 0.00 | 0.155 |
| Both | -0.03 | -0.06 | -0.01 | 0.052 | -0.04 | -0.07 | -0.01 | 0.024 |
| Grip strength | | | | | | | | |
| Neither | Reference | | | | | | | |
| Childhood | -0.02 | -0.04 | 0.00 | 0.098 | -0.03 | -0.05 | 0.00 | 0.061 |
| Adulthood | 0.05 | 0.03 | 0.07 | <0.001 | 0.05 | 0.03 | 0.06 | <0.001 |
| Both | 0.03 | 0.01 | 0.05 | 0.005 | 0.02 | 0.00 | 0.04 | 0.051 |

*Note:* CI = confidence interval. Model 1–adjusted for chronological age; Model 2–adjusted for chronological age, ethnicity, highest educational/professional qualification, annual gross household income and neighbourhood deprivation. *P*-values shown are corrected for multiple testing using the Benjamini–Hochberg procedure (across exposure levels, ageing markers and models). Sample sizes reported in Table S6.

#### 8. Sex-stratified associations childhood adversity (item-specific)

**Table S8.** Sex-stratified associations between childhood adversity items and ageing markers

| Females | Model 1 (adj. age) | | | | Model 2 (full adjustment) | | | |
|---|---|---|---|---|---|---|---|---|
| MileAge delta | *β* | 95% CI | | *p* | *β* | 95% CI | | *p* |
| Abuse, physical | 0.044 | 0.018 | 0.071 | 0.002 | 0.044 | 0.018 | 0.071 | 0.002 |
| Abuse, emotional | 0.054 | 0.028 | 0.080 | <0.001 | 0.052 | 0.026 | 0.078 | <0.001 |
| Abuse, sexual | 0.057 | 0.025 | 0.089 | 0.001 | 0.056 | 0.024 | 0.088 | 0.002 |
| Neglect, emotional | 0.006 | -0.036 | 0.048 | 0.836 | 0.007 | -0.035 | 0.049 | 0.830 |
| Neglect, physical | 0.020 | -0.004 | 0.044 | 0.149 | 0.018 | -0.005 | 0.042 | 0.189 |
| Metabolomic profile | | | | | | | | |
| Abuse, physical | 0.036 | -0.057 | 0.129 | 0.574 | 0.022 | -0.070 | 0.115 | 0.725 |
| Abuse, emotional | -0.013 | -0.105 | 0.079 | 0.836 | -0.029 | -0.120 | 0.062 | 0.644 |
| Abuse, sexual | 0.143 | 0.034 | 0.252 | 0.019 | 0.085 | -0.023 | 0.194 | 0.184 |
| Neglect, emotional | -0.020 | -0.174 | 0.135 | 0.846 | -0.076 | -0.230 | 0.078 | 0.445 |
| Neglect, physical | -0.007 | -0.089 | 0.076 | 0.902 | -0.040 | -0.123 | 0.042 | 0.445 |
| Frailty index | | | | | | | | |
| Abuse, physical | 0.324 | 0.308 | 0.339 | <0.001 | 0.303 | 0.287 | 0.318 | <0.001 |
| Abuse, emotional | 0.408 | 0.393 | 0.423 | <0.001 | 0.381 | 0.366 | 0.396 | <0.001 |
| Abuse, sexual | 0.321 | 0.302 | 0.340 | <0.001 | 0.297 | 0.279 | 0.316 | <0.001 |
| Neglect, emotional | 0.406 | 0.382 | 0.431 | <0.001 | 0.343 | 0.319 | 0.368 | <0.001 |
| Neglect, physical | 0.368 | 0.354 | 0.382 | <0.001 | 0.337 | 0.324 | 0.351 | <0.001 |
| Telomere length | | | | | | | | |
| Abuse, physical | 0.038 | 0.021 | 0.056 | <0.001 | 0.046 | 0.029 | 0.064 | <0.001 |
| Abuse, emotional | 0.021 | 0.003 | 0.038 | 0.034 | 0.026 | 0.009 | 0.044 | 0.006 |
| Abuse, sexual | 0.024 | 0.002 | 0.045 | 0.047 | 0.031 | 0.010 | 0.052 | 0.009 |
| Neglect, emotional | 0.045 | 0.018 | 0.073 | 0.003 | 0.044 | 0.017 | 0.072 | 0.004 |
| Neglect, physical | 0.017 | 0.001 | 0.032 | 0.062 | 0.019 | 0.003 | 0.034 | 0.036 |
| Grip strength | | | | | | | | |
| Abuse, physical | 0.009 | -0.001 | 0.018 | 0.107 | 0.003 | -0.007 | 0.012 | 0.666 |
| Abuse, emotional | 0.040 | 0.030 | 0.049 | <0.001 | 0.034 | 0.024 | 0.043 | <0.001 |
| Abuse, sexual | 0.006 | -0.005 | 0.018 | 0.419 | 0.000 | -0.012 | 0.011 | 0.968 |
| Neglect, emotional | 0.079 | 0.064 | 0.094 | <0.001 | 0.053 | 0.038 | 0.068 | <0.001 |
| Neglect, physical | 0.037 | 0.029 | 0.046 | <0.001 | 0.028 | 0.020 | 0.037 | <0.001 |
| Males | | | | | | | | |
| MileAge delta | | | | | | | | |
| Abuse, physical | 0.027 | 0.000 | 0.053 | 0.111 | 0.026 | -0.001 | 0.053 | 0.126 |
| Abuse, emotional | 0.020 | -0.013 | 0.053 | 0.391 | 0.017 | -0.015 | 0.050 | 0.453 |
| Abuse, sexual | -0.006 | -0.054 | 0.042 | 0.856 | -0.007 | -0.054 | 0.041 | 0.849 |
| Neglect, emotional | 0.020 | -0.029 | 0.068 | 0.537 | 0.013 | -0.036 | 0.063 | 0.687 |
| Neglect, physical | 0.019 | -0.008 | 0.045 | 0.294 | 0.016 | -0.011 | 0.043 | 0.392 |
| Metabolomic profile | | | | | | | | |
| Abuse, physical | 0.046 | -0.050 | 0.143 | 0.494 | 0.025 | -0.071 | 0.120 | 0.694 |
| Abuse, emotional | -0.015 | -0.135 | 0.105 | 0.856 | -0.041 | -0.160 | 0.078 | 0.596 |
| Abuse, sexual | -0.001 | -0.182 | 0.181 | 0.994 | -0.019 | -0.199 | 0.161 | 0.868 |
| Neglect, emotional | 0.163 | -0.021 | 0.347 | 0.168 | 0.126 | -0.058 | 0.310 | 0.315 |
| Neglect, physical | 0.074 | -0.022 | 0.169 | 0.248 | 0.055 | -0.040 | 0.150 | 0.403 |
| Frailty index | | | | | | | | |
| Abuse, physical | 0.255 | 0.239 | 0.271 | <0.001 | 0.230 | 0.214 | 0.246 | <0.001 |
| Abuse, emotional | 0.419 | 0.399 | 0.439 | <0.001 | 0.377 | 0.358 | 0.397 | <0.001 |
| Abuse, sexual | 0.258 | 0.229 | 0.286 | <0.001 | 0.242 | 0.214 | 0.270 | <0.001 |
| Neglect, emotional | 0.282 | 0.252 | 0.311 | <0.001 | 0.191 | 0.161 | 0.220 | <0.001 |
| Neglect, physical | 0.303 | 0.287 | 0.319 | <0.001 | 0.262 | 0.246 | 0.278 | <0.001 |
| Telomere length | | | | | | | | |

| | | | | | | | | |
|---|---|---|---|---|---|---|---|---|
| Abuse, physical | 0.026 | 0.007 | 0.044 | 0.016 | 0.026 | 0.007 | 0.044 | 0.017 |
| Abuse, emotional | 0.010 | -0.013 | 0.032 | 0.515 | 0.010 | -0.013 | 0.032 | 0.515 |
| Abuse, sexual | 0.012 | -0.020 | 0.044 | 0.582 | 0.014 | -0.018 | 0.046 | 0.515 |
| Neglect, emotional | 0.019 | -0.015 | 0.052 | 0.419 | 0.015 | -0.019 | 0.049 | 0.515 |
| Neglect, physical | -0.002 | -0.020 | 0.016 | 0.857 | -0.005 | -0.023 | 0.013 | 0.693 |
| Grip strength | | | | | | | | |
| Abuse, physical | -0.036 | -0.050 | -0.022 | <0.001 | -0.046 | -0.060 | -0.032 | <0.001 |
| Abuse, emotional | 0.064 | 0.047 | 0.081 | <0.001 | 0.038 | 0.020 | 0.055 | <0.001 |
| Abuse, sexual | 0.026 | 0.001 | 0.050 | 0.093 | 0.007 | -0.017 | 0.032 | 0.650 |
| Neglect, emotional | 0.103 | 0.077 | 0.129 | <0.001 | 0.052 | 0.026 | 0.078 | <0.001 |
| Neglect, physical | 0.036 | 0.022 | 0.050 | <0.001 | 0.016 | 0.002 | 0.030 | 0.055 |

*Note:* CI = confidence interval. Model 1–adjusted for chronological age; Model 2–adjusted for chronological age, ethnicity, highest educational/professional qualification, annual gross household income and neighbourhood deprivation. *P*-values shown are corrected for multiple testing using the Benjamini–Hochberg procedure (across childhood and adulthood exposures, ageing markers and models, separately for each sex). Sample sizes reported in Table S2.

#### 9. Sex-by-adversity interactions childhood adversity (item-specific)

**Table S9.** Sex interactions for associations between childhood adversity items and ageing markers

| | Model 1 (adj. age) | | | | Model 2 (full adjustment) | | | |
|---|---|---|---|---|---|---|---|---|
| MileAge delta | *β* | 95% CI | | *p* | *β* | 95% CI | | *p* |
| Abuse, physical | 0.02 | -0.02 | 0.05 | 0.534 | 0.02 | -0.02 | 0.05 | 0.534 |
| Abuse, emotional | 0.00 | -0.04 | 0.05 | 0.904 | 0.00 | -0.04 | 0.05 | 0.919 |
| Abuse, sexual | -0.05 | -0.11 | 0.00 | 0.182 | -0.05 | -0.11 | 0.00 | 0.186 |
| Neglect, emotional | -0.01 | -0.07 | 0.06 | 0.919 | -0.01 | -0.07 | 0.06 | 0.916 |
| Neglect, physical | 0.01 | -0.02 | 0.05 | 0.692 | 0.01 | -0.03 | 0.05 | 0.718 |
| Metabolomic profile | | | | | | | | |
| Abuse, physical | -0.01 | -0.14 | 0.13 | 0.939 | -0.01 | -0.15 | 0.12 | 0.895 |
| Abuse, emotional | -0.02 | -0.17 | 0.13 | 0.884 | -0.04 | -0.18 | 0.11 | 0.765 |
| Abuse, sexual | -0.14 | -0.36 | 0.07 | 0.348 | -0.11 | -0.32 | 0.10 | 0.481 |
| Neglect, emotional | 0.20 | -0.04 | 0.44 | 0.228 | 0.20 | -0.04 | 0.44 | 0.224 |
| Neglect, physical | 0.07 | -0.06 | 0.20 | 0.448 | 0.09 | -0.03 | 0.22 | 0.292 |
| Frailty index | | | | | | | | |
| Abuse, physical | -0.07 | -0.09 | -0.04 | <0.001 | -0.07 | -0.09 | -0.05 | <0.001 |
| Abuse, emotional | 0.01 | -0.01 | 0.04 | 0.448 | 0.00 | -0.03 | 0.02 | 0.939 |
| Abuse, sexual | -0.06 | -0.10 | -0.03 | 0.002 | -0.05 | -0.09 | -0.02 | 0.008 |
| Neglect, emotional | -0.13 | -0.16 | -0.09 | <0.001 | -0.14 | -0.18 | -0.11 | <0.001 |
| Neglect, physical | -0.06 | -0.09 | -0.04 | <0.001 | -0.07 | -0.09 | -0.05 | <0.001 |
| Telomere length | | | | | | | | |
| Abuse, physical | -0.02 | -0.05 | 0.00 | 0.208 | -0.03 | -0.05 | 0.00 | 0.084 |
| Abuse, emotional | -0.02 | -0.05 | 0.01 | 0.260 | -0.03 | -0.06 | 0.00 | 0.174 |
| Abuse, sexual | -0.01 | -0.05 | 0.02 | 0.626 | -0.02 | -0.06 | 0.02 | 0.534 |
| Neglect, emotional | -0.02 | -0.06 | 0.02 | 0.514 | -0.02 | -0.07 | 0.02 | 0.465 |
| Neglect, physical | -0.02 | -0.05 | 0.00 | 0.186 | -0.03 | -0.05 | 0.00 | 0.106 |
| Grip strength | | | | | | | | |
| Abuse, physical | -0.05 | -0.06 | -0.03 | <0.001 | -0.05 | -0.06 | -0.03 | <0.001 |
| Abuse, emotional | 0.02 | 0.00 | 0.04 | 0.059 | 0.02 | 0.00 | 0.03 | 0.224 |
| Abuse, sexual | 0.02 | -0.01 | 0.04 | 0.270 | 0.02 | 0.00 | 0.05 | 0.229 |
| Neglect, emotional | 0.02 | 0.00 | 0.05 | 0.224 | 0.01 | -0.02 | 0.04 | 0.627 |
| Neglect, physical | 0.00 | -0.02 | 0.01 | 0.885 | 0.00 | -0.02 | 0.01 | 0.708 |

*Note:* CI = confidence interval. Model 1–adjusted for chronological age; Model 2–adjusted for chronological age, ethnicity, highest educational/professional qualification, annual gross household income and neighbourhood deprivation. *P*-values shown are corrected for multiple testing using the Benjamini–Hochberg procedure (across childhood and adulthood exposures, ageing markers and models).

#### 10. Sex-stratified associations adulthood adversity (item-specific)

**Table S10.** Sex-stratified associations between adulthood adversity items and ageing markers

| Females | Model 1 (adj. age) | | | | Model 2 (full adjustment) | | | |
|---|---|---|---|---|---|---|---|---|
| MileAge delta | *β* | 95% CI | | *p* | *β* | 95% CI | | *p* |
| Abuse, physical | -0.003 | -0.030 | 0.024 | 0.859 | -0.007 | -0.034 | 0.020 | 0.711 |
| Abuse, emotional | 0.007 | -0.015 | 0.029 | 0.641 | 0.004 | -0.018 | 0.026 | 0.818 |
| Abuse, sexual | 0.011 | -0.023 | 0.044 | 0.644 | 0.006 | -0.027 | 0.040 | 0.802 |
| Neglect, emotional | 0.010 | -0.011 | 0.031 | 0.473 | 0.008 | -0.014 | 0.030 | 0.618 |
| Economic hardship | 0.004 | -0.023 | 0.031 | 0.836 | 0.000 | -0.027 | 0.028 | 0.975 |
| Metabolomic profile | | | | | | | | |
| Abuse, physical | -0.024 | -0.121 | 0.072 | 0.711 | -0.095 | -0.192 | 0.001 | 0.085 |
| Abuse, emotional | 0.003 | -0.073 | 0.078 | 0.958 | -0.022 | -0.097 | 0.054 | 0.675 |
| Abuse, sexual | -0.041 | -0.165 | 0.083 | 0.641 | -0.103 | -0.227 | 0.021 | 0.155 |
| Neglect, emotional | 0.098 | 0.023 | 0.172 | 0.019 | 0.043 | -0.032 | 0.119 | 0.373 |
| Economic hardship | 0.229 | 0.131 | 0.327 | <0.001 | 0.144 | 0.045 | 0.242 | 0.009 |
| Frailty index | | | | | | | | |
| Abuse, physical | 0.331 | 0.315 | 0.347 | <0.001 | 0.283 | 0.267 | 0.299 | <0.001 |
| Abuse, emotional | 0.345 | 0.332 | 0.358 | <0.001 | 0.319 | 0.307 | 0.332 | <0.001 |
| Abuse, sexual | 0.374 | 0.354 | 0.394 | <0.001 | 0.335 | 0.316 | 0.355 | <0.001 |
| Neglect, emotional | 0.238 | 0.225 | 0.250 | <0.001 | 0.174 | 0.161 | 0.187 | <0.001 |
| Economic hardship | 0.299 | 0.283 | 0.315 | <0.001 | 0.227 | 0.211 | 0.243 | <0.001 |
| Telomere length | | | | | | | | |
| Abuse, physical | 0.022 | 0.004 | 0.040 | 0.031 | 0.019 | 0.001 | 0.037 | 0.065 |
| Abuse, emotional | 0.018 | 0.003 | 0.032 | 0.028 | 0.017 | 0.002 | 0.031 | 0.043 |
| Abuse, sexual | 0.012 | -0.011 | 0.034 | 0.421 | 0.014 | -0.008 | 0.037 | 0.316 |
| Neglect, emotional | 0.007 | -0.008 | 0.021 | 0.483 | 0.007 | -0.007 | 0.022 | 0.444 |
| Economic hardship | 0.029 | 0.011 | 0.047 | 0.003 | 0.023 | 0.005 | 0.042 | 0.025 |
| Grip strength | | | | | | | | |
| Abuse, physical | 0.035 | 0.025 | 0.044 | <0.001 | 0.021 | 0.012 | 0.031 | <0.001 |
| Abuse, emotional | 0.030 | 0.022 | 0.038 | <0.001 | 0.025 | 0.017 | 0.032 | <0.001 |
| Abuse, sexual | 0.025 | 0.013 | 0.037 | <0.001 | 0.016 | 0.004 | 0.028 | 0.018 |
| Neglect, emotional | 0.050 | 0.042 | 0.057 | <0.001 | 0.027 | 0.019 | 0.035 | <0.001 |
| Economic hardship | 0.073 | 0.064 | 0.083 | <0.001 | 0.047 | 0.037 | 0.057 | <0.001 |
| Males | | | | | | | | |
| MileAge delta | | | | | | | | |
| Abuse, physical | -0.017 | -0.057 | 0.023 | 0.520 | -0.017 | -0.057 | 0.023 | 0.520 |
| Abuse, emotional | 0.001 | -0.029 | 0.031 | 0.959 | 0.002 | -0.028 | 0.032 | 0.924 |
| Abuse, sexual | -0.057 | -0.177 | 0.063 | 0.496 | -0.062 | -0.183 | 0.058 | 0.453 |
| Neglect, emotional | 0.008 | -0.015 | 0.032 | 0.588 | 0.001 | -0.023 | 0.026 | 0.924 |
| Economic hardship | 0.031 | -0.002 | 0.064 | 0.133 | 0.024 | -0.010 | 0.057 | 0.294 |
| Metabolomic profile | | | | | | | | |
| Abuse, physical | 0.064 | -0.082 | 0.210 | 0.515 | 0.033 | -0.113 | 0.178 | 0.735 |
| Abuse, emotional | 0.064 | -0.046 | 0.174 | 0.403 | 0.056 | -0.053 | 0.165 | 0.453 |
| Abuse, sexual | 0.515 | 0.034 | 0.997 | 0.084 | 0.422 | -0.054 | 0.898 | 0.168 |
| Neglect, emotional | 0.127 | 0.041 | 0.212 | 0.010 | 0.076 | -0.011 | 0.164 | 0.174 |
| Economic hardship | 0.143 | 0.015 | 0.270 | 0.068 | 0.081 | -0.047 | 0.209 | 0.368 |
| Frailty index | | | | | | | | |
| Abuse, physical | 0.253 | 0.229 | 0.277 | <0.001 | 0.230 | 0.206 | 0.254 | <0.001 |
| Abuse, emotional | 0.355 | 0.337 | 0.374 | <0.001 | 0.347 | 0.330 | 0.365 | <0.001 |
| Abuse, sexual | 0.573 | 0.499 | 0.646 | <0.001 | 0.494 | 0.423 | 0.566 | <0.001 |
| Neglect, emotional | 0.194 | 0.179 | 0.208 | <0.001 | 0.115 | 0.101 | 0.130 | <0.001 |
| Economic hardship | 0.277 | 0.257 | 0.297 | <0.001 | 0.181 | 0.161 | 0.201 | <0.001 |
| Telomere length | | | | | | | | |

| | | | | | | | | |
|---|---|---|---|---|---|---|---|---|
| Abuse, physical | 0.020 | -0.008 | 0.047 | 0.294 | 0.020 | -0.008 | 0.047 | 0.294 |
| Abuse, emotional | -0.009 | -0.029 | 0.012 | 0.524 | -0.008 | -0.029 | 0.013 | 0.547 |
| Abuse, sexual | 0.091 | 0.009 | 0.174 | 0.071 | 0.095 | 0.013 | 0.178 | 0.059 |
| Neglect, emotional | -0.010 | -0.026 | 0.006 | 0.377 | -0.014 | -0.031 | 0.002 | 0.186 |
| Economic hardship | 0.012 | -0.010 | 0.035 | 0.427 | 0.005 | -0.018 | 0.028 | 0.735 |
| Grip strength | | | | | | | | |
| Abuse, physical | 0.004 | -0.017 | 0.025 | 0.801 | -0.011 | -0.032 | 0.010 | 0.453 |
| Abuse, emotional | 0.028 | 0.012 | 0.044 | 0.002 | 0.018 | 0.003 | 0.034 | 0.055 |
| Abuse, sexual | 0.167 | 0.103 | 0.230 | <0.001 | 0.101 | 0.039 | 0.164 | 0.005 |
| Neglect, emotional | 0.116 | 0.103 | 0.128 | <0.001 | 0.075 | 0.062 | 0.088 | <0.001 |
| Economic hardship | 0.102 | 0.085 | 0.119 | <0.001 | 0.059 | 0.042 | 0.076 | <0.001 |

*Note:* CI = confidence interval. Model 1–adjusted for chronological age; Model 2–adjusted for chronological age, ethnicity, highest educational/professional qualification, annual gross household income and neighbourhood deprivation. *P*-values shown are corrected for multiple testing using the Benjamini–Hochberg procedure (across childhood and adulthood exposures, ageing markers and models, separately for each sex). Sample sizes reported in Table S5.

#### 11. Sex-by-adversity interactions adulthood adversity (item-specific)

**Table S11.** Sex interactions for associations between adulthood adversity items and ageing markers

| | Model 1 (adj. age) | | | | Model 2 (full adjustment) | | | |
|---|---|---|---|---|---|---|---|---|
| MileAge delta | *β* | 95% CI | | *p* | *β* | 95% CI | | *p* |
| Abuse, physical | 0.02 | -0.03 | 0.07 | 0.627 | 0.02 | -0.03 | 0.07 | 0.581 |
| Abuse, emotional | 0.02 | -0.01 | 0.06 | 0.361 | 0.03 | -0.01 | 0.06 | 0.314 |
| Abuse, sexual | -0.02 | -0.15 | 0.10 | 0.828 | -0.03 | -0.15 | 0.10 | 0.808 |
| Neglect, emotional | 0.00 | -0.03 | 0.03 | 0.939 | 0.00 | -0.03 | 0.03 | 0.952 |
| Economic hardship | 0.02 | -0.02 | 0.07 | 0.448 | 0.02 | -0.02 | 0.07 | 0.453 |
| Metabolomic profile | | | | | | | | |
| Abuse, physical | 0.08 | -0.10 | 0.25 | 0.534 | 0.12 | -0.05 | 0.30 | 0.314 |
| Abuse, emotional | 0.04 | -0.09 | 0.18 | 0.652 | 0.07 | -0.07 | 0.20 | 0.489 |
| Abuse, sexual | 0.54 | 0.04 | 1.04 | 0.113 | 0.51 | 0.02 | 1.00 | 0.132 |
| Neglect, emotional | 0.03 | -0.09 | 0.14 | 0.772 | 0.03 | -0.09 | 0.14 | 0.765 |
| Economic hardship | -0.09 | -0.25 | 0.07 | 0.454 | -0.07 | -0.23 | 0.09 | 0.534 |
| Frailty index | | | | | | | | |
| Abuse, physical | -0.08 | -0.10 | -0.05 | <0.001 | -0.05 | -0.08 | -0.02 | 0.007 |
| Abuse, emotional | 0.01 | -0.01 | 0.03 | 0.448 | 0.03 | 0.01 | 0.05 | 0.023 |
| Abuse, sexual | 0.20 | 0.13 | 0.28 | <0.001 | 0.17 | 0.09 | 0.24 | <0.001 |
| Neglect, emotional | -0.04 | -0.06 | -0.02 | <0.001 | -0.05 | -0.07 | -0.03 | <0.001 |
| Economic hardship | -0.02 | -0.05 | 0.00 | 0.224 | -0.04 | -0.06 | -0.01 | 0.021 |
| Telomere length | | | | | | | | |
| Abuse, physical | -0.01 | -0.04 | 0.02 | 0.626 | -0.01 | -0.04 | 0.02 | 0.708 |
| Abuse, emotional | -0.03 | -0.06 | -0.01 | 0.027 | -0.03 | -0.06 | -0.01 | 0.034 |
| Abuse, sexual | 0.06 | -0.02 | 0.15 | 0.288 | 0.07 | -0.02 | 0.15 | 0.264 |
| Neglect, emotional | -0.02 | -0.04 | 0.00 | 0.228 | -0.02 | -0.04 | 0.00 | 0.133 |
| Economic hardship | -0.02 | -0.05 | 0.01 | 0.448 | -0.02 | -0.05 | 0.01 | 0.364 |
| Grip strength | | | | | | | | |
| Abuse, physical | -0.03 | -0.05 | -0.01 | 0.015 | -0.02 | -0.04 | 0.00 | 0.141 |
| Abuse, emotional | 0.00 | -0.02 | 0.01 | 0.823 | 0.00 | -0.01 | 0.02 | 0.825 |
| Abuse, sexual | 0.14 | 0.08 | 0.20 | <0.001 | 0.11 | 0.06 | 0.17 | <0.001 |
| Neglect, emotional | 0.07 | 0.05 | 0.08 | <0.001 | 0.06 | 0.05 | 0.08 | <0.001 |
| Economic hardship | 0.03 | 0.01 | 0.05 | 0.014 | 0.02 | 0.00 | 0.04 | 0.116 |

*Note:* CI = confidence interval. Model 1–adjusted for chronological age; Model 2–adjusted for chronological age, ethnicity, highest educational/professional qualification, annual gross household income and neighbourhood deprivation. *P*-values shown are corrected for multiple testing using the Benjamini–Hochberg procedure (across childhood and adulthood exposures, ageing markers and models).
